# Travel-related Twenty-eight Days Cyclical Thrombosis and Subgroups of COVID-19 Cardiac Biomarker Data: Novel Review Strategy and Meta-analysis Method

**DOI:** 10.1101/2022.06.06.22275944

**Authors:** Keiichiro Kimoto, Munekazu Yamakuchi, Kazunori Takenouchi, Teruto Hashiguchi

## Abstract

**Backgrounds:** Strange controversies have remained in thrombosis-related fields (traveler’s and COVID-19-related thrombosis), although traveler’s thrombosis is well-known even among non-professionals as “economy class syndrome.” We hypothesized there might be something overlooked behind those strange situations.

**Methods:** Since ordinary review methods (e.g., systematic review or meta-analysis) had already been conducted, we focused on reviewing a “previously published” “chart.” Also, we developed a novel “review method” for the meta-regression analysis result. We applied those to some previously published and well-known data.

**Results:** We newly found an approximately 28 days cycle of thrombosis onset over several weeks after travel in a figure. Also, we found an eighteen-day cycle of thrombosis onset in another chart. In COVID-19 cardiovascular biomarker studies, we newly extracted subgroup patterns in a scatterplot (Troponin T and NT-proBNP) that applied simple linear regression analysis. Also, these subgroups had already appeared in the cardiomyopathy study.

**Conclusions:** Traveler’s thrombosis sometimes occurs over two months after leaving the risky in-flight environment. This phenomenon has been explained that the thrombus is formed in a cabin but dislodged after. However, from the cyclic patterns, explaining that the “high-risk period of thrombosis with Oral Contraceptive (OC) use initiation” coincided with “travel” is more reasonable (e.g., honeymoon and OC initiation). Regarding risk-benefit balance, it is conceivable that “spreading the risk” by starting the dosing away from the travel period is essential to ensure safer use because the in-flight environment may have a non-zero effect, and optimal care by a primary care physician (prescriber) is not available during the travel. In COVID-19, there seems to be a complex scatterplot structure that is unsuitable for usually used simple linear fitting. In a literature review, a pattern on a chart should be given more paying attention.

## Introduction

Traveler’s thrombosis is well-known even among non-professionals as “economy class syndrome.” However, controversial discussions have remained (**Supporting discussion 1: history of traveler’s thrombosis**)^1–17^. Also, the status of recent COVID-19-related thrombosis research seemed like that of the traveler’s thrombosis **(Supporting discussion 2: research situations of COVID-19-related thrombosis)**^18–23^. We hypothesized there might be something overlooked behind those strange situations.

However, trying ordinary review methods (e.g., systematic review or meta-analysis) again seemed impractical because those types of the study had already been conducted^2,6–10^. So, a novel strategy seemed to be needed.

Recently, there has been software that detects research fraud in the figures submitted for journals^24^, but it does not discover “overlooked findings.” Also, although raw data images, such as CT or MRI images, were analyzed, there is no awareness of new information extraction from the “previously published” “chart.” On the other hand, numerical data in tables within each piece of literature are usually evaluated in a meta-analysis. However, figures created by each researcher are generally not. Also, seeing the chart is more complex than we thought^25^. So, we focused on reviewing “figures.”

In addition, we focused on geometry. Although statistics usually have applied to medicine (e.g., p-value), approaching from a different direction seemed helpful when ordinary methods do not work.

We finally made our original strategy, which was extracting overlooked knowledge from the “previously published” “chart” by “geometrical viewpoint” **(Fig. S1, a-c)**.

Considering the above strategy, we focused on a figure that showed a result of meta-regression analysis **(Fig. 3, b)**(Chandra et al, *Ann Intern Med.* 2009;151(3):180-90., Figure 3)^10^. In this figure, we saw a geometrical shape (layered hyperbolic shapes) that seemed to be formed by two subgroups **(Fig. 3, Fig. S1, b)**. Also, we conceived a methodology, which was a manually executable kind of pattern recognition, searching layered hyperbolic patterns in a figure that shows a result of meta-regression analysis to detect latent sub-groups. Our basic idea is that a confidence interval length for an odds ratio is inversely proportional to the “potential patient population size” **(see Fig. S2, d-f)**.

In this paper, we report the results of our literature reviews based on our original strategy, including evaluating our methodological idea by fitting the curves derived from mathematical considerations **(the overall study framework is demonstrated in Fig. 2).**

## Methods

Geometrically, the edges of this hyperbolic shape are formed by a combination of an S-shaped curve and a U-shaped curve. We first focused only on the U-shaped curve.

The confidence interval length for the odds ratio is inverse proportional to the size of the potential patient population. However, to be precise, the ability of patient recruitment, in other words, the “sample size in each study,” also affects the confidence interval length. However, we speculated that the effect of the differences in sample size could not be a practical problem for two reasons.

The first reason is that stratified analysis, often performed in clinical studies, can be a force in reducing the differences. The second reason is that two types of boundary conditions in the hyperbolic shape fitting, which are the “S-curve” and the “error bar length,” reduce the degree of freedom in searching and increase the possibility of finding the desired shape.

Considering the numerical experiment **(Fig. 1, c)**, we decided on the equations for the S-shaped curve, the upper end of the confidence limit, and the lower end of the confidence limit (**Fig. 1, d**).

**Fig. 1.**
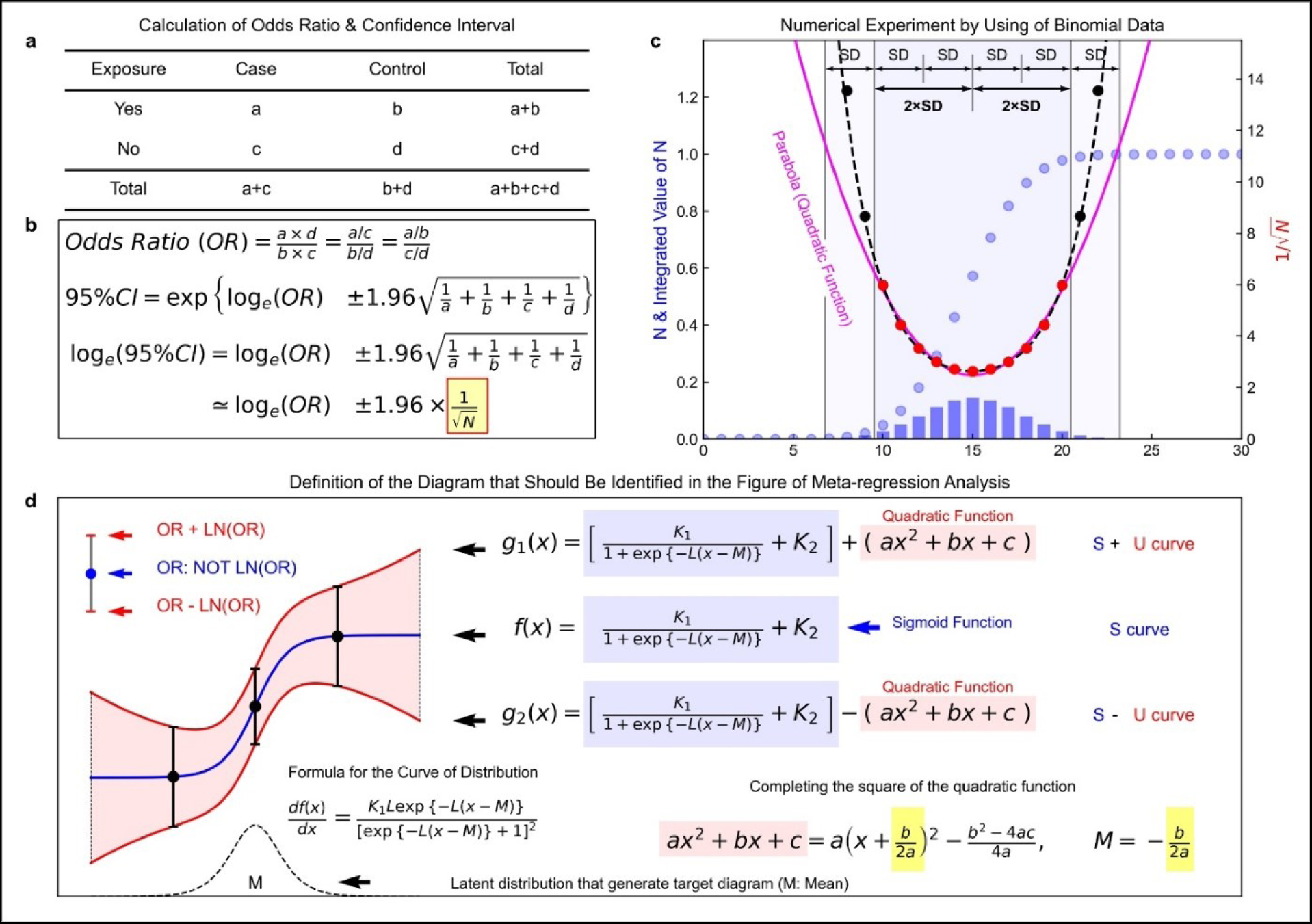
A hyperbolic shape formed by confidence limits. **a**, A two-way cross-tabulation (contingency table) for calculating an odds ratio (OR). **b**, Calculation of OR and its 95% confidence interval. **c**, Histogram of the data following a binomial distribution and a plot of the inverse values of the square roots. **d**, The mathematical formulas for the hyperbolic shape. Note that the OR is not logarithmic, and only the length of the error bar is logarithmic (see the upper left position of panel d). We generate binomial distribution data because normal distribution could be approximated by binominal distribution. Around the center of distribution (within ±2SD of the mean), the parabola was a good fit for the points calculated from the simulated data. Also, from around the over ±2SD to ±3SD range, the parabola is positioned below the points. Although a catenary curve (dashed curve) is more likely suitable for fit, the number of patients who fall into the categories at both ends is slightly larger than the expected number. Usually, data placed over the range of ±3 SD from the distribution center are summarized as “greater than” or “less than.” For example, in the case of 10 hours is the center of the distribution, the time category might be following sets such as “less than” 5 hours, 6-8 hours, 9-11 hours, 12-14 hours, and “more than” 15 hours. The value of 1 over root N (1/sqrt(N)) becomes smaller. So, using a parabola seemed to be based on the reality of the data handling method in the medical and biological fields.

The whole process, including driving the formulas, data review, regression analysis, and tools for the analysis, is described in the supplementary information (**Supporting information: Method details**).

## Results

### 1) Analysis 1A (dataset: Chandra et al)^10^

Before redrawing the figures of the meta-regression analysis, we reviewed the cited four studies (Martinelli et al, 2003; Parkin et al, 2006; Cannegieter et al, 2006; Kuipers et al, 2007)^1,3,5,26^. The main objective of Chandra et al’s research^10^ was estimating a risk, but the main of our analysis is detecting the subgroups. So, we are considering the possibility of some problems with inclusion in our study, even if there were no problems in Chandra et al’s research.

In this assessment, two study data (Martinelli et al, 2003; Cannegieter et al, 2006)^1,3^ were judged inappropriate and excluded from our analysis. Cannegieter et al and Martinelli et al used a unique research design, which uses the patient’s partner as a control (Martinelli et al: friend and partner). In the introduction, we described that although the properties of each study (the ability of patient recruitment, sample size) disrupt the effect of the size of the potential patient population, the stratified analysis could reduce the disruption. However, the impact of adding a unique study design was unclear.

Additionally, the odds ratio reported by Parkin et al^26^ was suspected to be affected by the misplacement of the cells in the cross table for calculating the odds ratio. Still, qualitatively, it could be used **(see Fig. 3, Table S1, Table S2, and Supporting information: Method details)**.

In the residual dataset (Parkin et al, 2006 & Kuipers et al, 2007)^5,26^, layered hyperbolic patterns appeared, and applied regression analysis with the above three formulas. The inflection points of the S-curves (center of distributions) were 7.1 hours and 11.8 hours, respectively **(Fig. 3, b).**

Additionally, it seemed that S-curve could be fitted to the group of points reported by Cannegieter et al and Martinelli et al, respectively. Also, those of inflection points would position at positions of less than 10 hours and more than 10 hours. This result seems consistent with the two hyperbolic shapes fitting **(Fig. 3, b, orange, and green dots)**.

Notably, we observed an overlooked cyclic pattern of thrombosis onset in the figure reported by Cannegieter et al during the above data review process. The cycle period was 27.4 days **(Fig. 4, a-e)**. The wave of about 28 days consisted of a gradually decaying wave and a monotonically decreasing function **(Fig. 4, e)**. The wave disappeared after about three months (90 days). The monotonically decreasing part estimated that it continued for over 90 days after the travel **(Fig. 4, f)**. Cannegieter et al did not mention this wave pattern.

### 2) Analysis 1B (dataset: Philbrick et al)^7^

Considering that only a few pieces of data were available for the first analysis, we performed a similar analysis using another dataset gathered by Philbrick et al^7^ for a systematic review. As a preparation, we conducted a data review to confirm eligibility for our regression analysis **(see Fig. 2)**. The dataset did not contain risk ratio data. In this case, hyperbolic shapes could not be fitted (for the details, see **Supporting information: Method details**). So, only S-curves were applied. This analysis found two S-curves in the stratified datasets by pulmonary embolism (PE) and deep vein thrombosis (DVT). The inflection point of PE was about 9.2 hours, and the point of DVT was about 12.1 hours, respectively **(Fig. S3, b)**, which were similar to the first analysis.

**Fig. 2.**
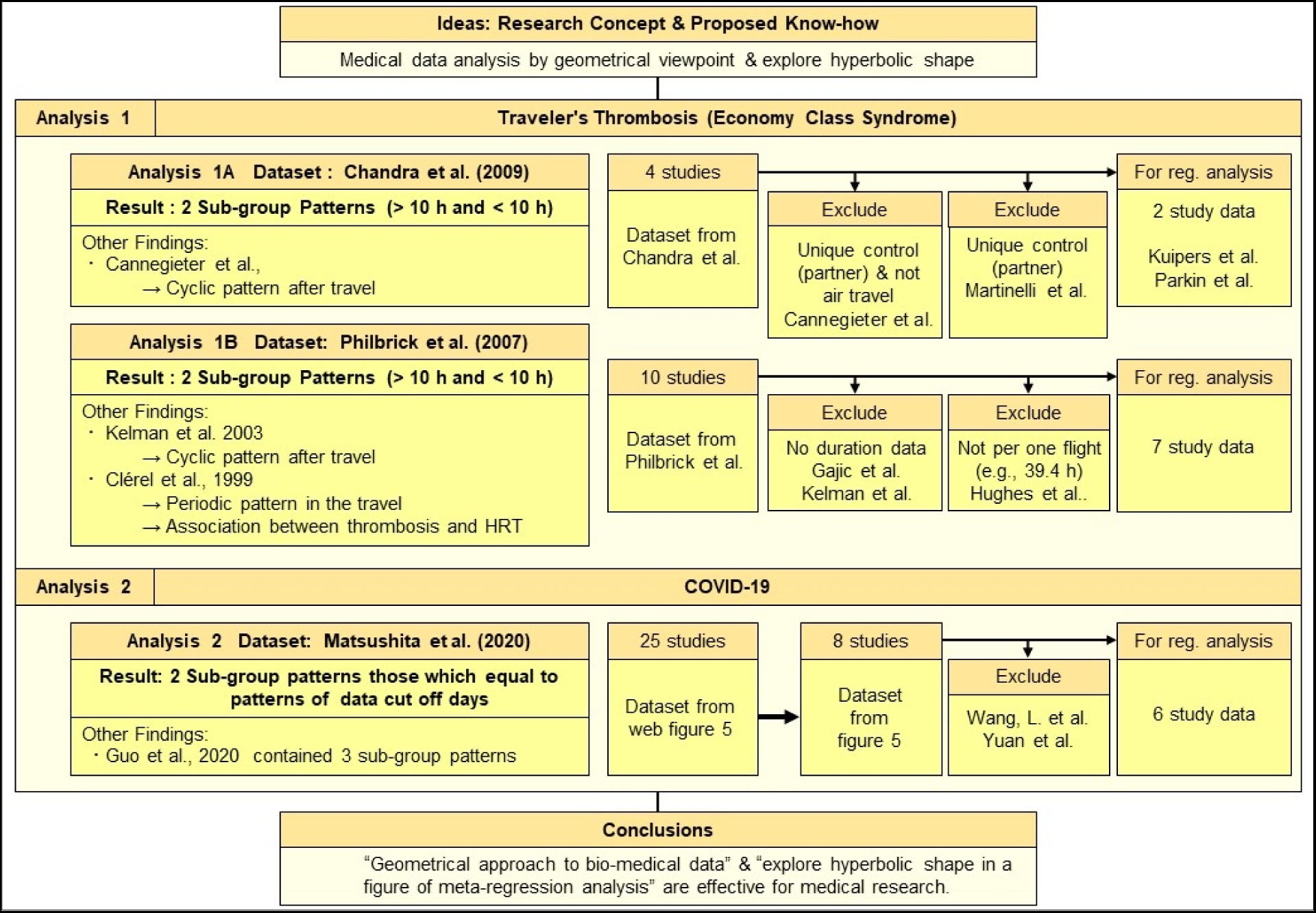
Overall framework of this study. We applied our concepts to travelers’ thrombosis and COVID-19. These were similar in research history (see **Supporting discussion 1: history of traveler’s thrombosis** & **Supporting discussion 2: research situations of COVID-19-related thrombosis**). In the case of traveler’s thrombosis, we analyzed a dataset collected by Chandra et al and Philbrick et al^7,10^. In the case of COVID-19, we analyzed a dataset collected by Matsushita et al^10^. Flow charts show data accept or reject flows. To avoid duplicating data, Matsushita et al selected only eight studies data (25 studies included in initial web Figure 5)^36^.

Additionally, we found that unrecognized periodic pattern in the data reported by Clérel and Caillard^27^ during the data review, which was suspected to be the effect of circadian rhythm **(Fig. S4, c).**

We also found a cyclic pattern after travel in the data reported by Kelman et al^28^. However, the period of the wave was 17.9 days, and the wave was less clear than the wave observed in the data reported by Cannegieter et al. In the waveform, there was a raw-risk period just before 90 days. Also, in this data, there was an uptrend **(Fig. 5)**.

Furthermore, we found a correlation between the stepped stages of increasing PE patients and the issuance of the guidelines^29–32^ for preventive hormone replacement therapy (HRT) to a menopause woman in a figure reported by Clérel and Caillard^27^ **(Fig. S5)**. Especially the thrombosis onset in Paris airports has increased since the late 1990s **(Fig. S5)**.

### 3) Analysis 1B-related additional analysis

We performed the trend analysis of the literature to assess the generalization on the increasing number of patients at Paris airports reported by Clérel & Caillard^27^ **(Fig. S5)**. We searched the literature in PubMed (keywords: “Economy Class Syndrome,” “Traveler’s Thrombosis,” and “Travel-related thrombosis”). The number of references increases rapidly after the 2000s **(Fig. S6)**.

The difference between the above result of trend analysis and the result in the data reported by Clérel and Caillard^27^ can be explained by the difference between publication day and onset day. So, there is no practical difference between the two results.

The literature search found that two articles (Symington & Stack, 1977, Cruickshank, Gorlin, & Jennett, 1988)^33,34^ contained each patient’s details. We already obtained each patient’s data from the other two articles in analyses 1A and 1B (Clérel & Caillard, 1999, Parkin et al, 2006)^26,27^. So, we confirm changing the sex ratio using those data to assess the effect of HRT. We found that the rate of women in the PE data gathered from the above four studies^26,27,33,34^ gradually increased but was most pronounced during the late 1990s **(Table S3).**

We also conducted a literature review targeting early literature because we strongly questioned the “economy class syndrome” concept from our findings in Analysis 1A and 1B. We found that the word appeared in Symington & Stack, 1977^33^. However, judging from the citation history checked on the Europe PMC website (https://europepmc.org/), the article by Cruickshank, Gorlin, & Jennett, 1988^34^ might influence the word as evangelists. The paper was a “personal paper,” a type of article category of the Lancet, and the authors prepared case reports of their own experiences.

Dr. Cruickshank was middle-aged^34^. Usually, the mid-life crisis is concerned around this range of age. Dr. Jennett was a dean of medicine at Glasgow^35^. Generally, heavy psychological stress based on heavy social responsibility is concerned. So, we hypothesized that men’s life event-related psychological stress could be one of the risk factors **(see Fig. S7, b).**

To assess our hypothesis, we merged data from the above four studies^26,27,33,34^ and made a bar chart for 1-year. The onset of PE was concentrated in certain age stages, such as around 30, the 40s, late 50s, and early 70s **(Fig. S7, e)**.

We performed the same analysis in the women’s data except for data reported by Clérel & Caillard^27^. Although the peak range was 65 to 70 in the data of Clérel & Caillard^27^, PE was concentrated in the 40s to 50s (menopausal stage) in this analysis **(Fig. S8, d)**.

Additionally, we found an inverted-U-shaped (mountainous) relationship in the scatter plot: age on the horizontal axis and air travel time on the vertical axis **(Fig. S9)**. However, it was possible to fit a straight line as an up-right linear relationship or a down-right linear relationship **(Fig. S9, a, and Fig. S10, a, c)**.

These analyses were also performed on the female data reported by Clérel & Caillard^27^, and we found a U-shaped relationship **(Fig. S9, c)**.

### 4) Analysis 2 (dataset: COVID-19)^36^

We decided to apply our ideas to the current complex COVID-19 pandemic problem to find some overlooked things because there was some controversy^18–23^ about COVID-19-related thrombosis.

As a note, we described that stratified is necessary for the introduction section, but clinical studies of COVID-19 did not perform it. However, Stratification seemed unnecessary because the size of the studies was not huge and roughly the same due to no mega trial during the pandemic.

One of us (KK) searched for the layered hyperbolic pattern using the search engine Google and extracted a figure reported by Matsushita et al^36^ **(Fig. S11, a-c)**. After evaluating eligibility **(see Fig. 2)**, patterns that allowed to be fitted hyperbolic shapes appeared **(Fig. S11, d-f, & g-i)**.

Considering the cause of these patterns, we found the matching between grouping the data by hyperbolic patterns and grouping by data cut-off days **(Fig. S12)**. We calculated weighted averages of age by grouping. In the earlier days group, non-severe was 46.5, and severe was 57.7, which were middle age (age 45-64). In the late date group, non-severe was 40.9, and severe was 68.6, which were mature age (age 25-44) and elderly (age > 65), respectively.

Surprisingly, we found unrecognized patterns during the data review process in a figure reported by Guo et al.^37^, which showed a relationship between N-terminal pro-brain natriuretic peptide (NT-proBNP) and Troponin T (TnT). There were three clusters of subgroup patterns, and each cluster could be fitted parabola **(Fig. 6, a, b, Fig. S13, a, b & Fig. S14, a, d, e-h)**.

The third subgroup pattern was similar to ST-elevating myocardial infarction in the figure report by Budnik et al^38^. That study was a comparative study between Takotsubo cardiomyopathy (also known as stress cardiomyopathy or broken heart syndrome) and ST-elevating myocardial infarction. This tilted parabola appeared in another study on COVID-19 patients **(Fig. S13, c)**^39^, and the pattern had already appeared in other studies^40,41^ before the pandemic **(Fig. S14, b & c)**. However, patterns in COVID-19 patients were not recognized by each author.

Additionally, In the case of dropping points to the horizontal axis (troponin axis), the histogram on the troponin axis results in bimodality. The above results matched the patterns reported by other studies **(Fig. S13, b, d & e)**^39,42,43^.

Furthermore, by mounting the data of high-sensitivity C-Reactive protein (hsCRP) onto the third subgroup in the data reported by Guo et al^37^ and constructing a parabolic cylinder, we found a pattern that might further be subdivided into two subgroups on the side surface of this parabolic cylinder **(Fig. 6, c, d, & Fig. S15)**.

## Discussion

### 1) Traveler’s thrombosis

#### a. Hyperbolic patterns

In traveler’s thrombosis, we obtained consistent results from two analyses (Analysis 1A: 7.1 hours and 11.8 hours, Analysis 1B: 9.2 hours and 12.1 hours) **(Fig. 3, b, Fig. S3, b)**. The discrepancy between the value of 7.1 hours and 9.2 hours seems to be explained by the miscalculation found in the data review process **(see Table S1, Table S2, and Supporting information: Method details)**.

**Fig. 3.**
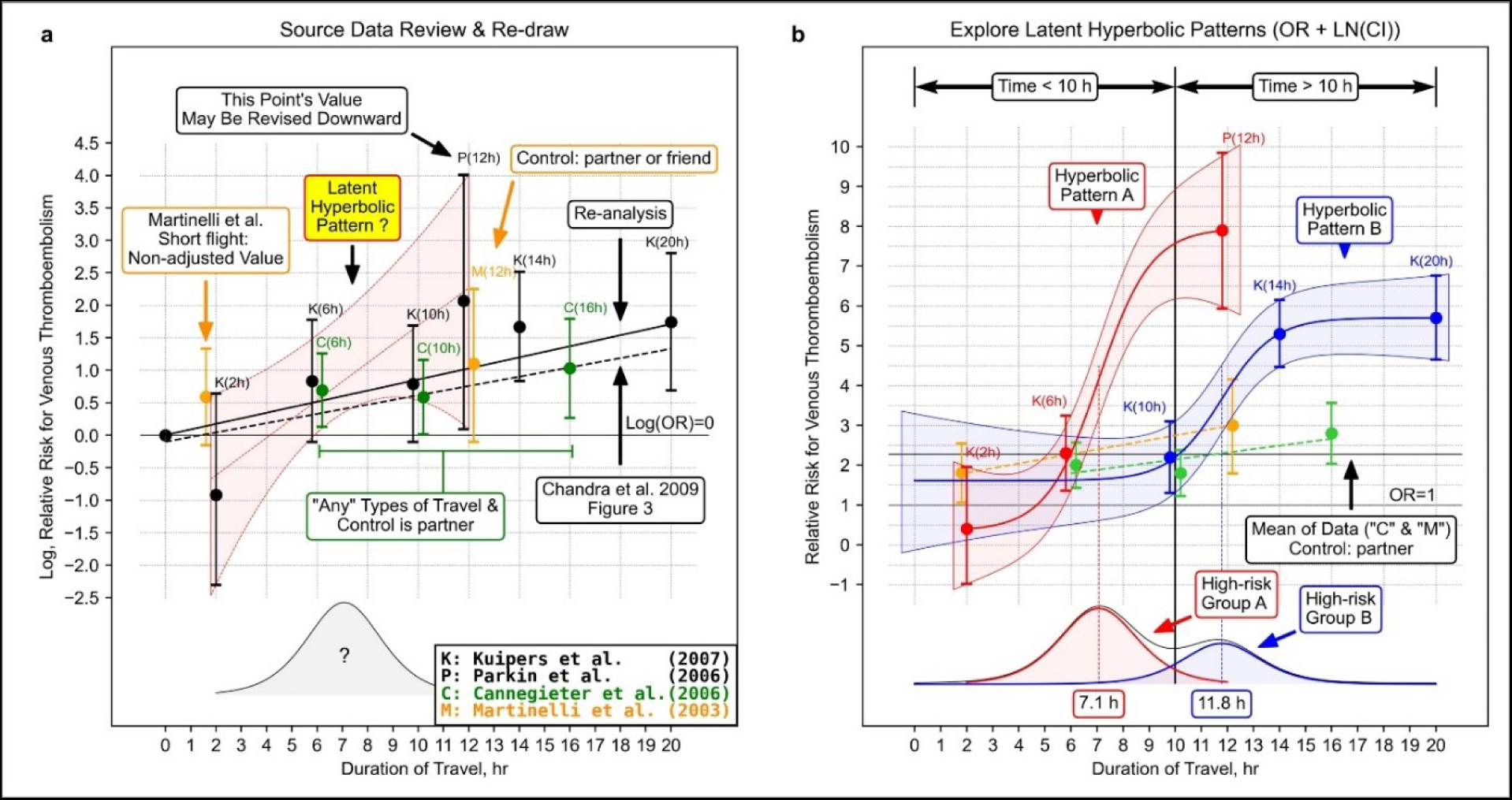
Re-analysis based on the proposed ideas. **a**, Data review and meta-regression analysis using the same method as Chandra et al. *Ann Intern Med*. 2009; 151(3):180-190. Figure 3^10^. We added information from original studies and our hypothesis to the previously published form, such as latent distribution. This figure is regarded as a re-drawn figure from Chandra D, Parisini E, Mozaffarian D. Meta-analysis: travel and risk for venous thromboembolism. *Ann Intern Med*. 2009 Aug 4;151(3):180-90. doi: 10.7326/0003-4819-151-3-200908040-00129. Epub 2009 Jul 6. © 2009 American College of Physicians. Adapted with permission. The original figure has been shown on the American College of Physicians website, which links to PubMedR○ (https://pubmed.ncbi.nlm.nih.gov/19581633/). **b**, grouping the data and hyperbolic shape fitting. The dots represent the odds ratio with error bars of the confidence interval are the adjusted odds ratio described in the four studies cited by Chandra et al. Since Cannegieter et al did not describe values of confidence intervals, we calculated the values from the case-count data. We also calculated the odds ratio at the time point of short travel in the research reported by Martinelli et al. Chandra et al did not display it might be due to the no adjusted odds ratio initially described. Our result of a meta-regression analysis was slightly different from the straight line drawn by Chandra et al might be due to the software used by Chandra et al (STATA) being different from the one we used (R and R package “metafor”) or they might conduct a meta-regression analysis reversing the front head and the front side of the cross-tabulation. It was impossible to fit S-shaped curves for the points excluded from the search for hyperbolic shapes (Martinelli et al & Cannegieter et al) due to the small number of data or the form of the point sequence. However, if we manually fit S-curves, the two inflection points would appear close to the positions of the hyperbolic shapes’ inflection points.

Our results implied two S-curves. Also, we hypothesized two high-risk periods and two types of high-risk groups. In those groups, “factor V Leiden paradox”^44^ may have to be considered **(Supporting discussion 3: two high-risk periods and two types of high-risk groups?)**.

#### b. Cyclic patterns

Another noteworthy point is the cyclic patterns in the data reported by Cannegieter et al^3^ **(Fig. 4).** In Cannegieter et al^3^, the cyclic pattern disappeared around three months (90 days), consistent with the high-risk period in oral contraceptive (OC) use was the first three months^45^. Also, there is multi-phase OC in the category of OC. Therefore, it is conceivable that the 28 days cycle OC is attributable to the cyclic pattern.

**Fig. 4.**
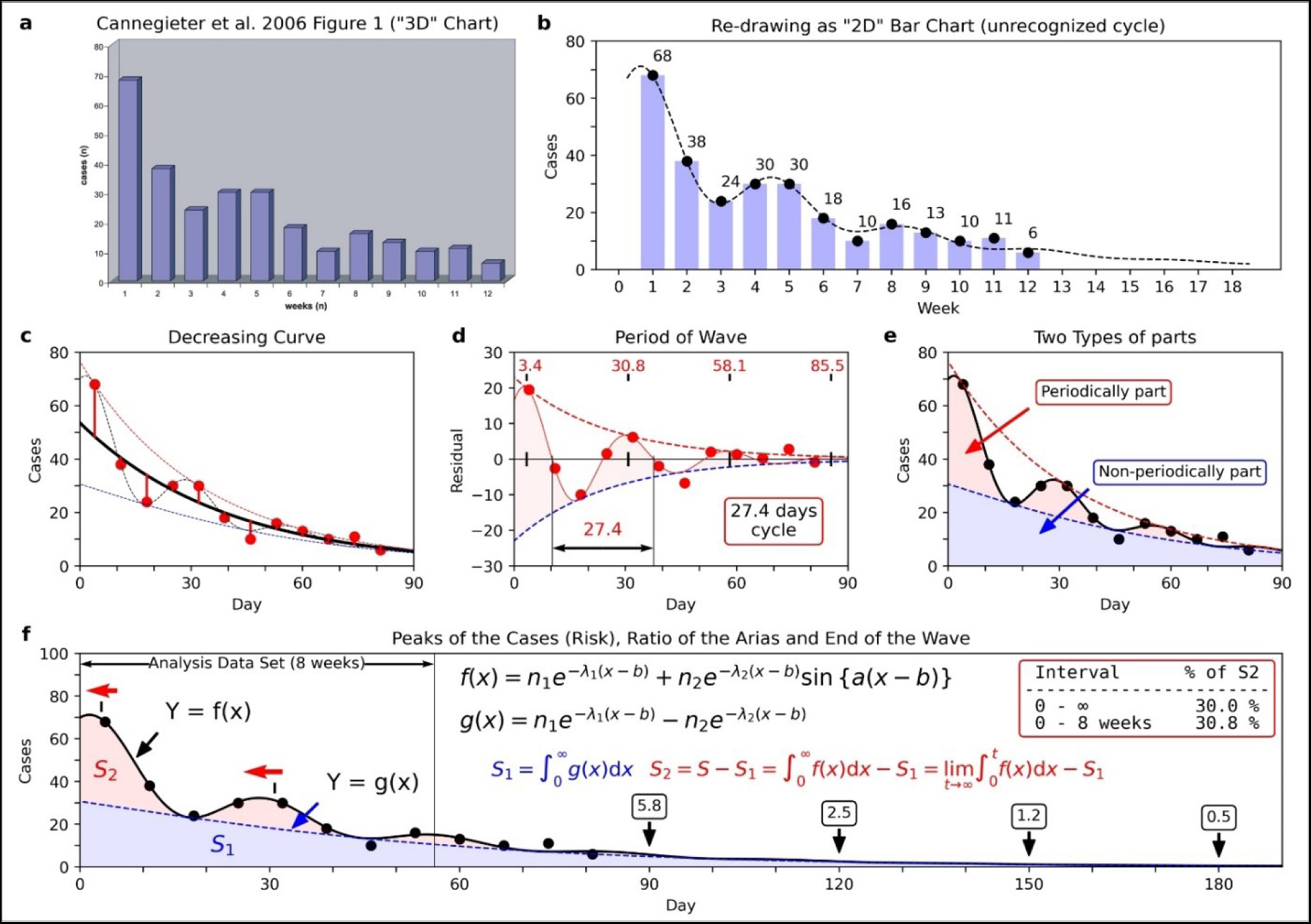
Unrecognized cyclic pattern of thrombosis onset after travel^3^. **a**, A bar graph showing the relationship between weeks after travel and the number of thrombosis onset in a figure reported by Cannegieter et al^3^. **b**, Re-expressing as a two-dimensional bar graph avoiding the three-dimensional representation. **c**, Applying a damped wave by non-linear regression analysis. **d**, Extraction of damped wave part by subtracting the monotonic decrease function. **e**, Dividing into 2 sub-parts areas by the envelopes of the damped wave that touch the lower parts of the wave. **f**, Enlarged and added explanation of the ratio of the area. Cannegieter et al^3^ used the data up to week eight. The ratio of sub-parts 2 to the total number of patients (ratio of S2 to the area of S1 + S2) was 30.0%. In the integration for the infinite interval, it was 30.8%. Peak shifts of the wave (peak position of the wave shifted from the original to the other) appeared when comparing panels d and f due to putting by regression curve located in the center (c + d). Panel a was re-used from Cannegieter et al. *PLoS Med*. 2006; 3(8):e307. Figure 1. https://www.ncbi.nlm.nih.gov/labs/pmc/articles/PMC1551914/figure/pmed-0030307-g001/ Copyright © 2006 Cannegieter et al. Creative Commons Attribution License. In 2006, the Creative Commons Attribution 2.0 Generic, License (CC BY 2.0) was available. https://creativecommons.org/licenses/by/2.0/

Additionally, the raw-risk period just before 90 days in the data reported by Kelman et al^28^ **(Fig. 5)**. This raw-risk period could be explained that extended-use type OC, which has a planned drug withdrawal, such as 84 active days and seven placebo days^46^. Also, the uptrend in the data by Kelman et al might be due to depot agent type OC administrated in 90 days cycle, which injects drugs periodically after decreasing the drugs in the body.

**Fig. 5.**
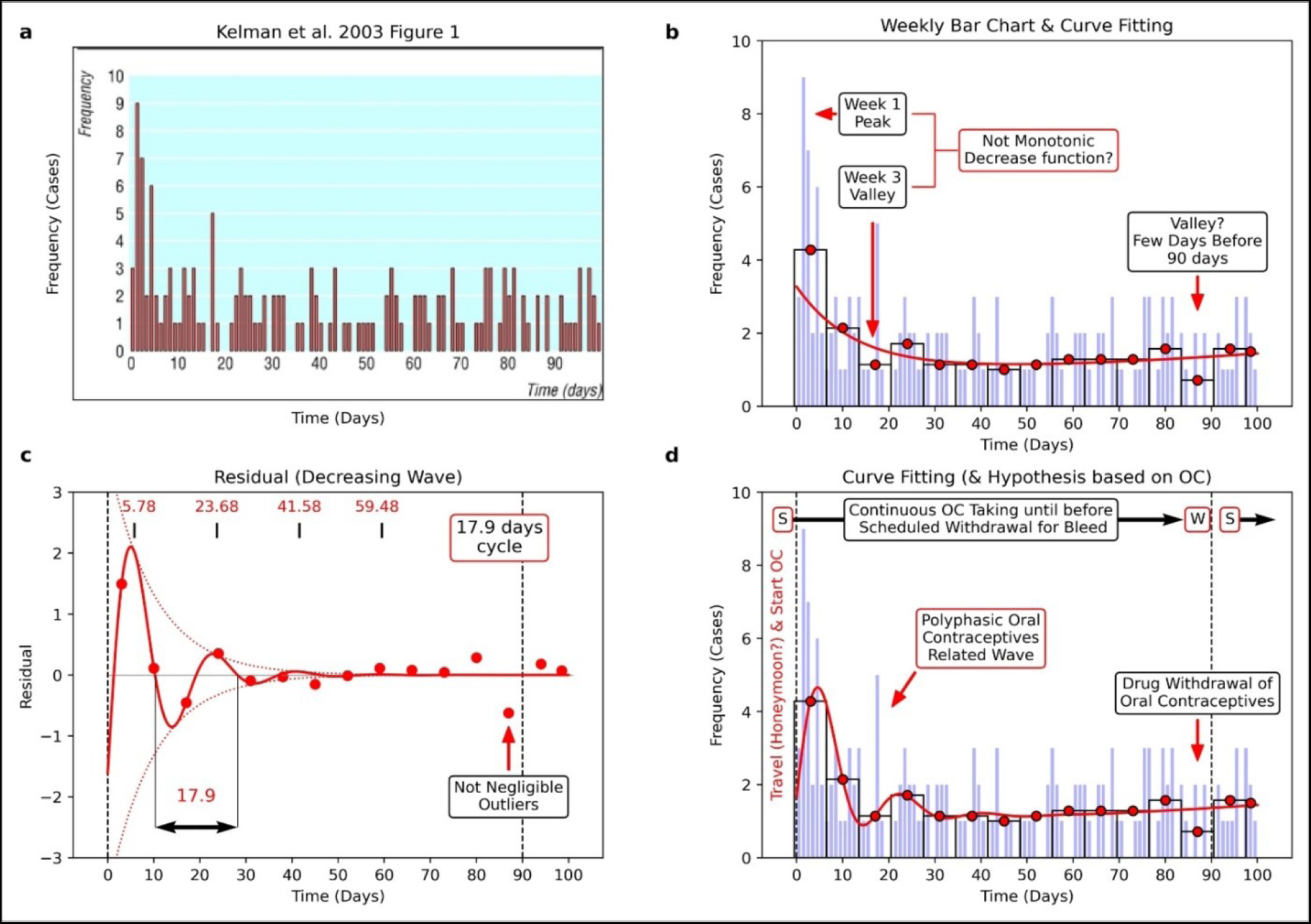
Cyclic pattern of thrombosis onset appeared in a figure by Kelman et al^28^. **a**, A thrombosis onset distribution reported by Kelman et al^28^. **b**, The regression curve is located in the center of all average points (the points express the average of thrombosis onset during seven days). **c**, Curve fitting to the residual data of the regression curve in panel b. **d**, Application of damped wave function and adding interpretation of the results assuming some women started taking oral contraceptives (OC) in the timing of travel. The observed cyclic pattern was relatively unclear than Cannegieter et al^3^ **(see** Fig. 4). Noteworthy, the thrombosis onset downed in the days before 90 days, and it seems to be the scheduled withdrawal period of OC use. This figure was re-used and re-drawn from Kelman et al. *BMJ*. 2003; 327(7423):1072. Figure 1 https://www.bmj.com/content/327/7423/1072#F1 Copyright © 2003 BMJ Publishing Group Ltd. All rights reserved. The BMJ permission team thankfully confirms this figure adaptation. Also, we obtained permission to re-use.

Based on the above results, those types of OC use (multi-phase type, extended-use type, and depot agent type) might be triggered by travel.

#### c. Difference of the periods and patterns

Interestingly, there was a difference between the period of cyclic patterns (Cannegieter et al: about 28 days, Kelman et al: about 18 days). Also, the wave in the data reported by Cannegieter et al was clear, but the wave in the data reported by Kelman et al was not clear.

Regarding the halfway cycle (about 18 days) in Kelman et al, it is notable that a combination of daily estrogen dosing and 10–14 days of progestogen^47^ is used for HRT. Also, in 1999, Clérel and Caillard mentioned that OC and HRT might affect thrombosis after travel^27^.

It is also notable that thrombosis onset in the data reported by Cannegieter et al was “March 1999-March 2000” (only most late 1990s to early 2000)^3^. Thrombosis onset in the data reported by Kelman et al was “1981-1999” (including the whole of the 1990s)^28^.

Therefore, the difference in waves can be explained as follows: the halfway cycle (17.9 days) in Kelman et al was a mixture of the 28 days OC cycle and a type of HRT cycle. However, only OC users might remain in the data reported by Cannegieter et al after the issue of the HERS study in 1998^31^.

#### d. Specific age in man?

Analysis 1B-related additional analysis suggested that the development of travel-related thrombosis occurs at specific ages **(Fig. S7)**. On the other hand, Kelman et al reported the incidence of VTE per 100,000 passengers^28^. In this data, the incidence of VTE sharply increased after age 40, and it was a temporary decrease in the early 60s. Although it should be noted that these results are not limited to men, they are consistent with our results.

We also found that the relationship between male age and travel duration seems to be inverted-U-shaped (mountainous) **(Fig. S9 & Fig. S10)**. Regarding this result, men’s life event-related psychological stress may be one of the risk factors.

#### e. In-flight environment?

There has been a widespread belief that in-flight environments cause thrombosis. Also, the phenomenon of the onset of thrombosis after over a few weeks to two months since the travel has been explained, such as the development of blood clots occurring on a cabin, but the dislodge of a thrombus occurs after^34^. Also, although OC-related thrombosis had already been known, OC was recognized as not a substantial cause but one of the risk (aggravating) factors^1^.

Other researchers might realize other possibilities changed the name of thrombosis after travel from “traveler’s thrombosis” to “travel-related thrombosis,” but they could not show evidence.

However, the above cyclic patterns of thrombosis onset may suggest that the disease concept of traveler’s thrombosis (travel-related thrombosis) should be changed. From the cyclic patterns, it is more reasonable to explain that the “travel” and “high-risk period of thrombosis with OC use (within three months from starting OC)” coincided with the travel (e.g., honeymoon and starting birth control OC).

#### f. Life-event-related thrombosis

Recently, Er et al reported a few cases of venous thromboembolism (VTE) incidence during the COVID-19 lockdown^48^. However, all patients had inter mediate-high risk pulmonary embolisms. Therefore, the influence of the in-flight environment may be less than we had previously thought. From a comprehensive view, “traveler’s thrombosis” or “travel-related thrombosis” may require to be regarded as “life-event-related thrombosis.”

Even if a substantial risk factor is not the in-flight environment, it is not neglectable at the present research stage. Our thought that starting OC or HRT was triggered by travel suggests that usage based on risk diversification is important to avoid overlapping risks of drug and in-flight environment. Also, well-planned may allow more safe use of OC and HRT. Additionally, for men, perhaps we should focus more on “stress reduction.” (**Supporting discussion 4: for more discussion on Traveler’s thrombosis in Women and Men**).

### 2) COVID-19

In applying our ideas for COVID-19, the dataset grouping by Matsushita et al^36^ matched the grouping by data cut-off dates **(Fig. S12)**. Age structure diverged from middle age to mature and elderly. Middle-aged people (age 45-64) might enter the workforce for manual labor treating fish containers, mature people (age 25-44) might want to select IT jobs, and elderly people (age > 65) might stay in the house. So, our results were consistent with the hypothesis that COVID-19 spread from the seafood market.

In our data review process **(Supporting discussion 5: linear regression analysis)**, our analysis of the data reported by Guo et al^37^ showed three clusters of subgroups and formed the tilted parabola. Also, the patterns matched the other studies **(Fig. 6, a, b, Fig. S13 & Fig. S14)**^38–43^. Considering the study by Budnik et al^38^, those subgroups may have different biological mechanisms. Also, we found patterns on the side surface of this parabolic cylinder **(Fig. 6, c, d, & Fig. S15)**.

**Fig. 6.**
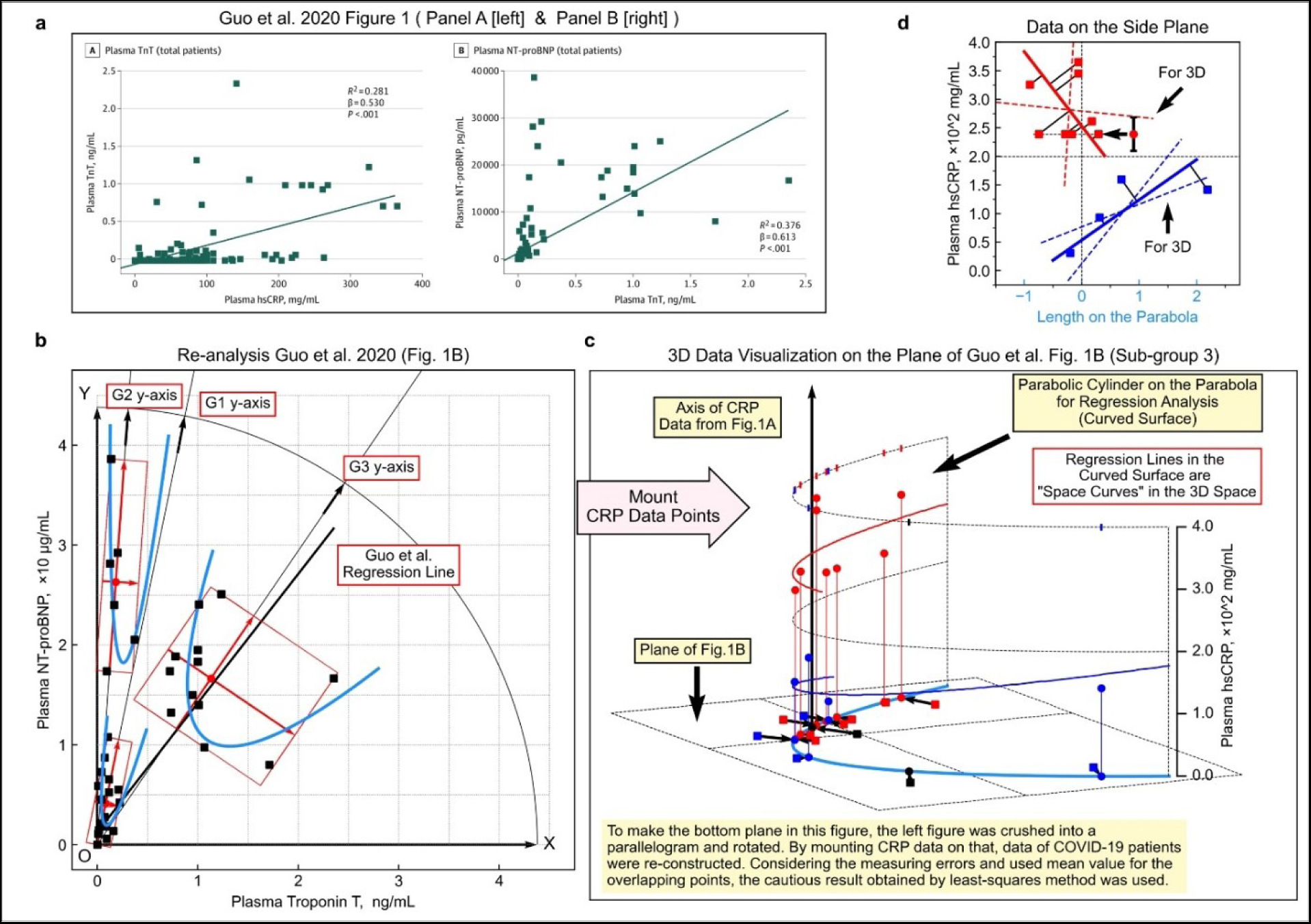
Parabola shape patterns in the figure reported by Guo et al^37^. **a**, Relationship between high sensitive C-reactive protein (hsCRP) and cardiac troponin T (TnT) in COVID-19 patients (left) and the relationship between cardiac troponin T (TnT) and N-terminal pro-brain natriuretic peptide (NT-proBNP) (right). **b**, Data points from the right side of panel a and fitting parabola. c, Three-dimensional data visualization was constructed by mounting the value of hsCRP onto panel b. **d**, Linear regression analysis on the side surface of the parabolic cylinder in panel **c**. The points were replaced with their average if the values could not be determined due to overlapping (the points indicated by the left-pointing arrow and the error bar, which are the average and the range of values, respectively). Panel a was re-used from Guo T et al. *JAMA Cardiol*. 2020; 5(7):811-818. Figure 1 https://www.ncbi.nlm.nih.gov/labs/pmc/articles/PMC7101506/figure/hoi200026f1/ Copyright © 2020 Guo T et al. *JAMA Cardiology*. Creative Commons Attribution License (CC-BY). https://creativecommons.org/licenses/by/4.0/

Meisel et al^49^ mentioned that the CRP to troponin ratio (CRP/troponin) could serve to differentiate between myopericarditis and acute myocardial ischemia (AMI), although not the study on COVID-19 patients. In COVID-19, Caro-Codón et al reported interesting behavior of CRP^42^. A meta-analysis by Lagunas-Rangel reported that the lymphocyte-to-C-reactive protein ratio (LCR) level, which was not a simple measurement value but a ratio, might be related to an inflammatory process^50^.

The above studies might imply complex data structures in 2-dimensional and 3-dimensional scatter plots consisting of biomarker values, and our findings may serve the cardiac biomarkers’ research field.

### 3) Conclusions

From the consistent results of Analysis 1A and 1B and the explainable result of analysis 2, it is conceivable that our strategy and method were effective.

We could not judge whether our findings are the root cause of the remaining controversies in thrombosis research. However, judging from many findings, when conducting a literature review, although paying attention to the reproducibility of a pattern in charts is needed to avoid arbitrary discussions, more serious consideration should be given to a pattern on a chart that has been ignored as a noisy variation.

## Acknowledgments

One of the authors, Keiichiro Kimoto, appreciates the kindful encouragement of Hideo Yoshioka, MEcon., who is in charge of the Data Strategy Research Institute representative.

## Statement of our intention for data analysis results on previous published articles

In addition to general acknowledgments, to clarify our stance, we mention this statement of intention for results. In this article, we pointed out many overlooked information and errors. However, we have no intention to attack previous works because our analysis results owing to their original works, including original research articles, reported valuable data, and writings of meta-analysis synthesized valuable datasets. Since just a scientist had better reconfirm the previous studies with no preconceptions and being grateful to those studies’ researchers, we carefully reviewed the data reported by previous studies. We highly respect previous works intending to solve medical issues.

## Transparency declaration

This study was not required ethical approval because the study used ONLY two types of openly available human data. Firstly, this study performed meta-analyses as secondary analyses. The source of the data was described in the method section. Second, this study analyzed measured values from figures in previous published scientific papers.

All the figures were appropriately treated with careful attention to the copyright issues to protect the integrity of the science. The sources of the original articles were described in the figure legend following the copyright holder’s instructions. Also, we submit copyright details in another file to a journal that we submit this manuscript file to explain additional information.

This study suggests a new review strategy and meta-analysis method for exploratory data analysis and reports some epidemiological findings. So, there were no applicable guidelines (e.g., Preferred Reporting Items for Systematic reviews and Meta-Analyses, PRISMA). However, this study carefully follows the usage of the statistical analysis method, and the whole process is precisely described in the method details section and **Fig. 2**.

This study was not received financial support from any funding.

## Data availability statement

We analyzed clinical data published by other studies (third parties). Used data is identified by indicated information of citation (reference numbers and list of references). The corresponding author responds to inquiries in the case of measured values from published figures requested by reviewers or readers.

## Code availability statement

Correspondence author (KK) can respond to inquiries for the corresponding author’s email address on offering the Python source code and spreadsheet software files for statistical analysis. Also, we have already uploaded our Python programs for the following GitHub repository to keep traceability perfectly.

https://github.com/Keiichiro-KIMOTO-Kagoshima-University/Travel-related_Twenty-eight_Days_Cyclical_Thrombosis_and_Subgroups_of_COVID-19_Cardiac_Biomarker_Dat

## Notes

### Competing Interest Statement

Munekazu Yamakuchi, Kazunori Takenouchi, and Teruto Hashiguchi have completed the Unified Competing Interest form and declare: no support from any organization for the submitted work; no financial relationships with any organizations that might have an interest in the submitted work in the previous three years, no other relationships or activities that could appear to have influenced the submitted work. Keiichiro Kimoto has been in charge of external advisor for the Data Strategy Research Institute but received no financial support for this study.

### Funding Statement

This study did not receive any funding.

### Author Declarations

This study was not required ethical approval because the study used ONLY two types of openly available human data. Firstly, this study performed meta-analyses as secondary analyses. The source of the data was described in the method section. Second, this study analyzed measured values from figures in previous published scientific papers. All the figures were appropriately treated with careful attention to the copyright issues to protect the integrity of the science. The sources of the original articles were described in the figure legend following the copyright holder's instructions. Also, we submit copyright details in another file to a journal that we submit this manuscript file to explain additional information. This study suggests a new review strategy and meta-analysis method for exploratory data analysis and reports some epidemiological findings. So, there were no applicable guidelines (e.g., Preferred Reporting Items for Systematic reviews and Meta-Analyses, PRISMA). However, this study carefully follows the usage of the statistical analysis method, and the whole process is precisely described in the method details section and Fig. 2. This study was not received financial support from any funding.

### Summary of Updates

Although this revision is basically just an adjustment of the number of figures and words to the submission rules of a journal, and the changes in the whole text are minor, the critical sentences were added in the abstract section. In our manuscript, we have mentioned that the central issue in travel-related thrombosis patients was an initiation of Oral Contraceptives (OC) and Hormone Replacement Therapy (HRT) associated with travel. However, regarding the impact on readers of the periodic fluctuation (wave) diagrams included in our study results, we had thought that enough consideration should be paid to making our manuscript to avoid superfluous warning inhibits a person who truly needs access to OC and HRT. Also, since the manuscript on the preprint server is not peer-reviewed but is easily accessible for non-specialists online, we wanted to be as careful as possible. So, we inserted sentences that mention how to proceed with safer usage of OC and HRT in our manuscript (discussion section). However, we had not included this discussion in the abstract section, which is the most conspicuous part. So, we decided to add considerations in the discussion section of the previous manuscript to the abstract area in the new manuscript, and we thought that the revised manuscript should be uploaded on the preprint server even though the changes in the whole text are minor.

