## Supporting Information for "Travel-related Twenty-eight Days Cyclical Thrombosis and Subgroups of COVID-19 Cardiac Biomarker Data: Novel Review Strategy and Meta-analysis Method"

This PDF file includes:

- Supporting figures & figure legends (Figures S1 to S15)
- Supporting tables & table legends (Tables S1 to S3)
- Method details
- Supplementary discussions
- SI References

**NOTE**

**Especially in the "Method details" section, we pointed out many overlooked information and errors. However, we have no intention to attack previous works because our analysis results owing to their original works, including original research articles, reported valuable data, and writings of meta-analysis synthesized valuable datasets. We highly respect previous works intending to solve medical issues.**

### 1. Supporting figures & figure legends

**Fig. S1.**

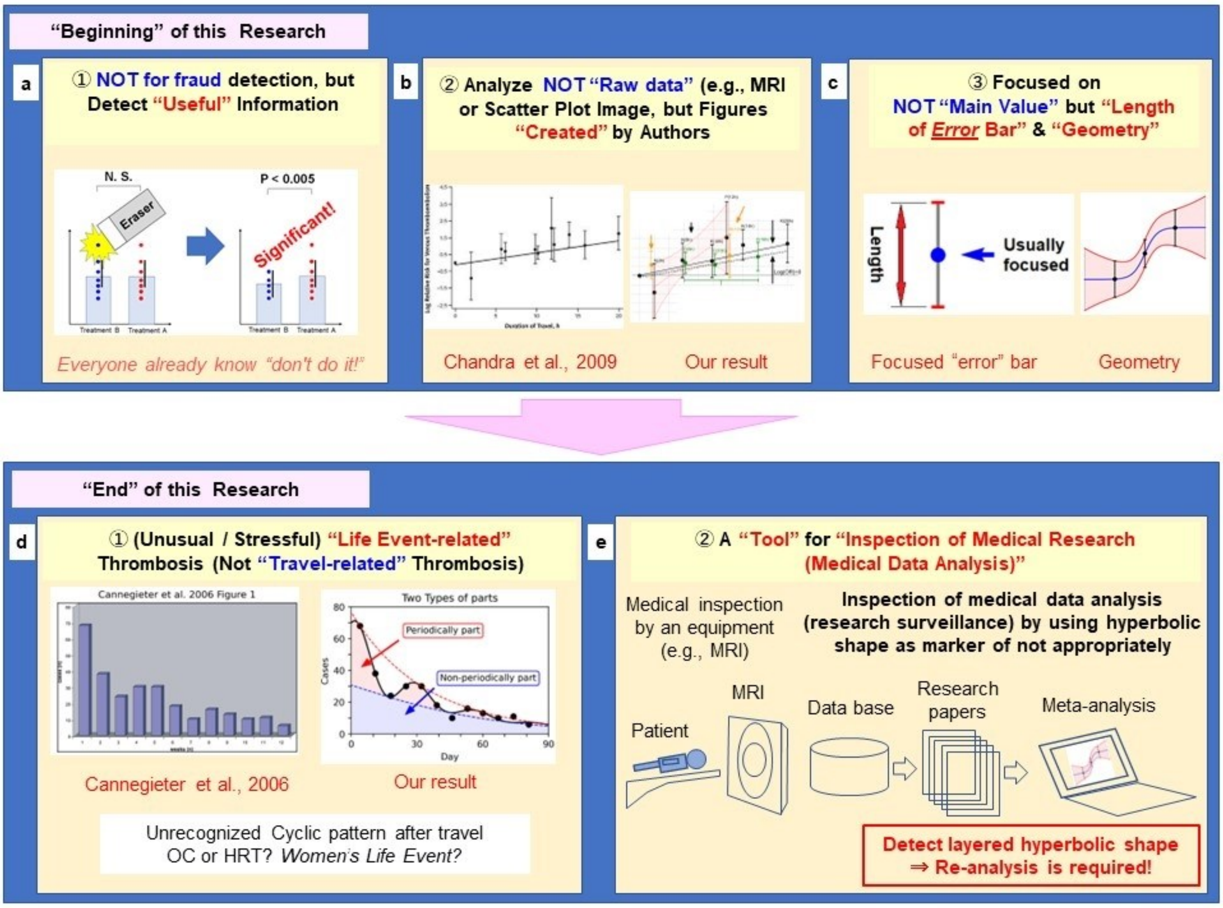


**Fig. S1. Illustration of our concepts at the beginning and the end of this study.**

**a**, Extracting does not research fraudulent but useful findings that had been missed. **b**, Finding latent useful findings not from image data such as MRI or X-ray images (a kind of raw data) but from charts created by each researcher. **c**, Finding latent useful findings from not value itself is usually interesting, but a geometric consideration of the length of the error bars of the confidence intervals, which used to evaluate the main value. **d**, a new concept, "life event-related thrombosis," replacing the concept of travel-related thrombosis. **e**, Monitoring whether clinical research society executes appropriate data analysis, as an application of the subgroup search methodology proposed in this study. In panel b, on those figures re-used and re-drawn are derived from Chandra D, Parisini E, Mozaffarian D. Meta-analysis: travel and risk for venous thromboembolism. *Ann Intern Med*. 2009 Aug 4;151(3):180-90.

doi: 10.7326/0003-4819-151-3-200908040-00129. Epub 2009 Jul 6. © 2009 American College of Physicians. Adapted with permission. The original figure has been shown on the American College of Physicians website, which links to PubMed®(https://pubmed.ncbi.nlm.nih.gov/19581633/). In panel d, those figure are re-used or re- drowned from Cannegieter et al. *PLoS Med*. 2006; 3(8):e307. Figure 1. https://www.ncbi.nlm.nih.gov/labs/pmc/articles/PMC1551914/figure/pmed-0030307-g001/ Copyright © 2006 Cannegieter et al. Creative Commons Attribution License. In 2006, the Creative Commons Attribution 2.0 Generic, License (CC BY 2.0) was available. https://creativecommons.org/licenses/by/2.0/

**Fig. S2.**

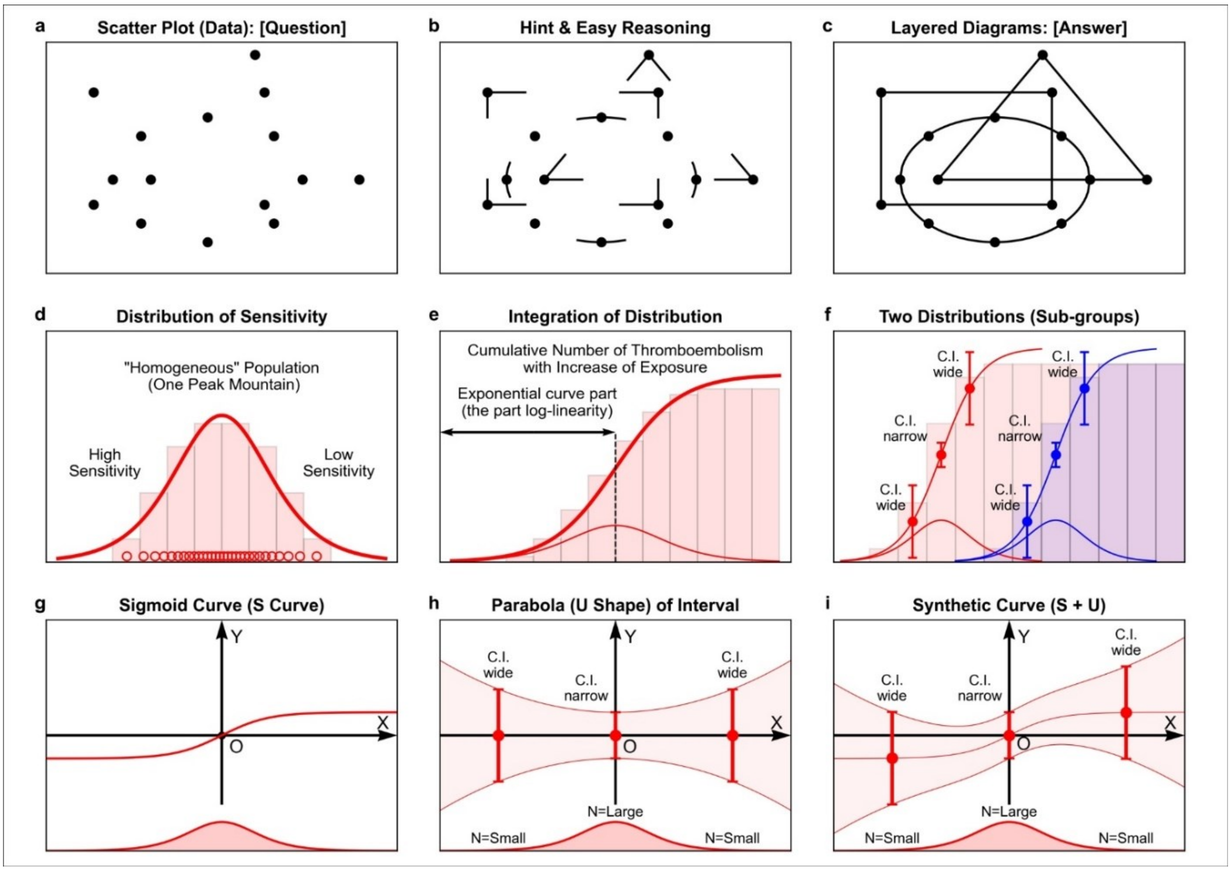


**Fig. S2. Finding shapes using a "key" to search and "latent patient population" onset with a strength of risk.**

**a**, The apex points of the circle, triangle, and quadrilateral. **b**, The apex points with a part of the edge close to the points. **c**, Complete shapes of triangles, quadrilaterals, and circles. **d**, Schematic representation of a homogeneous latent patient population. **e**, Schematic representation of the cumulative number of cases. **f**, Schematic depiction of two subgroups. **g**, A schematic representation of the S-curve formed by the cumulative distribution. **h**, A schematic representation of the width of the confidence interval. **i**, A schematic representation of S-curve plus width of the confidence interval. Although it is difficult to find the overlapped shapes from the only apex points, it is easy to arrive at the correct answer if we obtain some "key" or "hints" (panels a-c). Our idea, searching subgroups focusing on the layered hyperbolic shape to find, is similar to finding latent patterns using some key or hint. The number of cases increases with the integration of exposure to risk factors. There may be two sub-group if there are two types of origin of S-curves in the x-axis that represent the strength of risk (panels d-f).

**Fig. S3.**

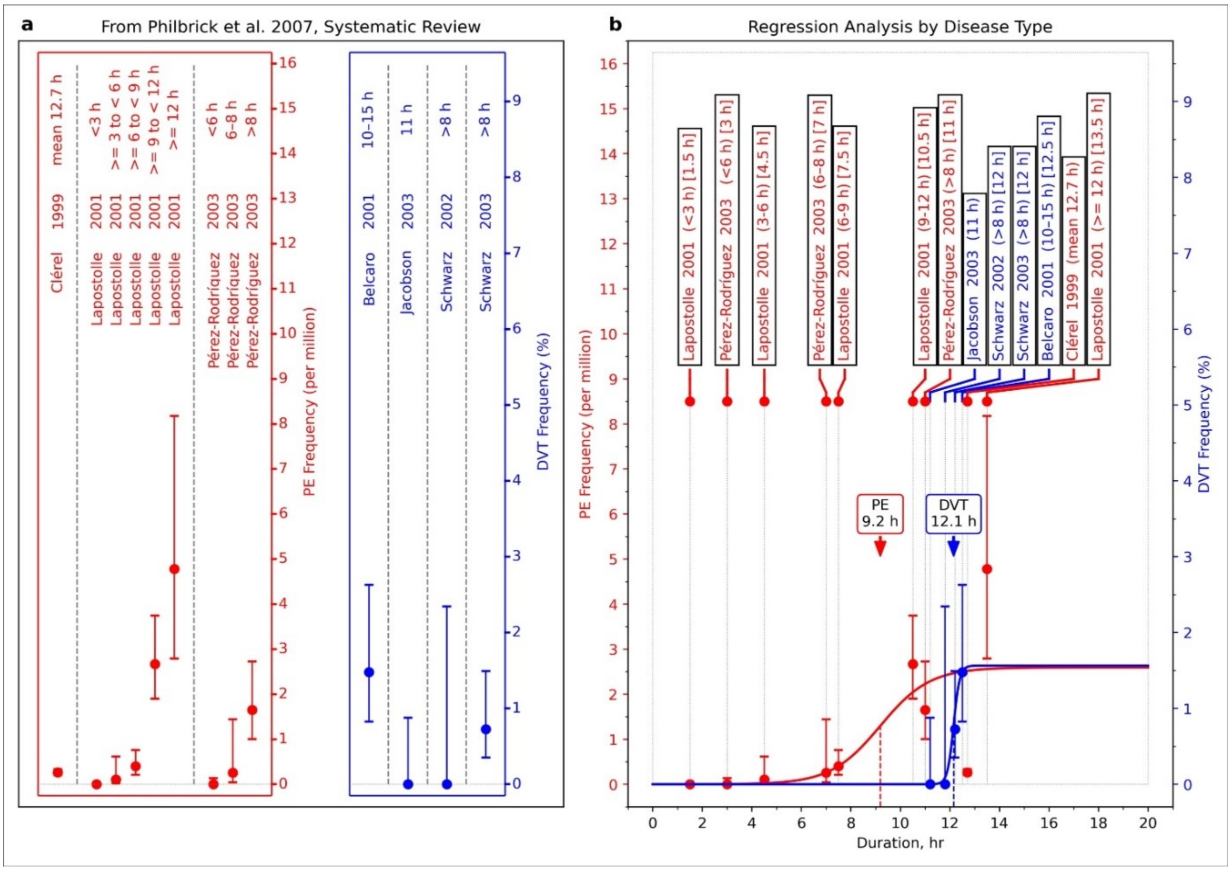


**Fig. S3. Curve fitting to the dataset reported by Philbrick et al[7].**

**a**, Data stratification by Pulmonary Embolism (PE) and Deep Vein Thrombosis (DVT). **b**, Application of S-shaped curve by regression analysis to the stratified data. The value in the bracket (panel b) is the point of time converted from the time category (see **Supporting information: Method details**). Philbrick et al[7] reported the result of a systematic review with a only table (list).

**Fig. S4.**

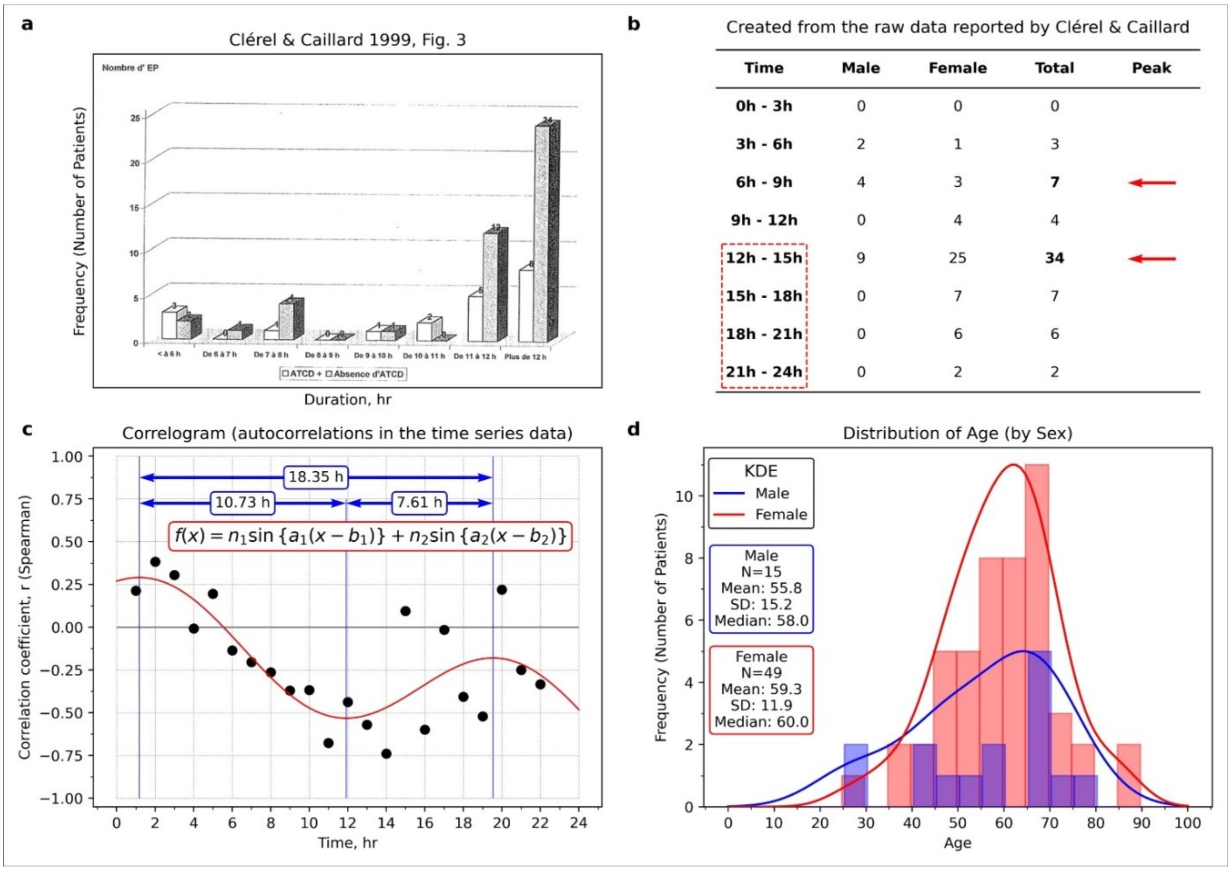


**Fig. S4. Re-analysis of the data in Table 1 reported by Clérel & Caillard[27].**

**a**, A figure made by Clérel & Caillard[27] showed a relationship between travel time and the thrombosis onset in the case of stratification by medical history of thrombosis. **b**, The relationship between time and thrombosis (prepared from Table 1 reported by Clérel & Caillard[27]). **c**, Correlogram (prepared from Table 1 reported by Clérel & Caillard[27]). **d**, Age distribution by sex (made from Table 1 reported by Clérel & Caillard[27]). As shown in panel a, Clérel & Caillard[27] summarized all the data for 12 hours or more, but there were two peaks (see panel b). In panel c, there was a periodic pattern. Panel a reproduced from Clérel & Caillard. Syndrome thrombo-embolique de la station assise prolongée et vols de longue durée: l'expérience du Service Médical d'Urgence d'Aéroports De Paris. *Bull Acad Natl Med*. 1999; 183(5):985-997. discussion 997-1001. Figure 3. Copyright © 1999 Elsevier Masson SAS. All rights reserved. Académie Nationale de Médecine.

**Fig. S5.**

**a**

Clérel & Caillard 1999, Fig. 1.

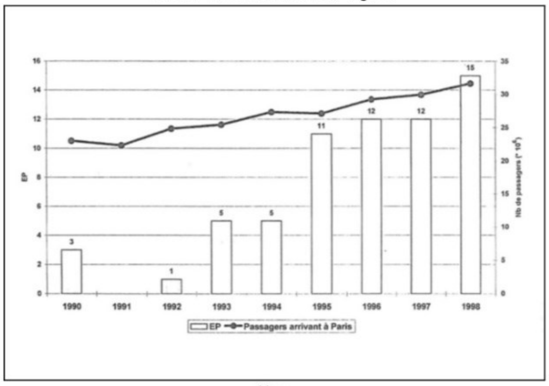


Year

**c**

History of Hormone Replacement Therapy (Recommendations to Concerns of Safety)

**Timeline Table 1: Years 1992–1997 (Recommendations of Hormone Replacement Therapy, HRT)**

| Year | Month | Type of Article | Journal (Citation) | Recommendation |
|---|---|---|---|---|
| 1992 | Dec | ACP Guideline | Ann Intern Med. 1992;117(12):1038-41. | Consider for coronary disease |
| 1995 | Jul | AHA Guideline | J Am Coll Cardiol. 1995;26(1):292-4. | Mentioned risk reduction interventions |
| 1997 | May | AHA Guideline | Circulation. 1997;95(9):2329-31. | Consider HRT in postmenopausal women |
| 1997 | Jun | NHS Cohort | N Engl J Med. 1997;336(25):1769-75. | Mortality: HRT user < non-user |

**Timeline Table 2: Years 1998–2002 (Expressing Concerns on the Safety of Hormone Replacement Therapy, HRT)**

| Year | Month | Type of Article | Journal (Citation) |
|---|---|---|---|
| 1998 | Aug | HERS Randomized Clinical Trial | AMA. 1998;280(7):605-13. |
| 1999 | May | AHA/ACC scientific statement | J Am Coll Cardiol. 1999;33(6):1751-5. |
| 2001 | Jul | AHA Guideline | Circulation. 2001;104(4):499-503. |
| 2002 | Jul | HERS Randomized Clinical Trial Follow-up (HERS II) | AMA. 2002;288(1):58-66. |
| 2002 | Jul | Women's Health Initiative (WHI) Randomized Clinical Trial | JAMA. 2002;288(3):321-33. |

**Abbreviations:**
ACP: American College of Physicians, AHA: American Heart Association, NHS: Nurses' Health Study
HERS: Heart and Estrogen/progestin Replacement Study, ACC: American College of Cardiology

**Fig. S5. Thrombosis reported by Clérel & Caillard [27] & our novel annotations.**

**a**, Clérel & Caillard mentioned that "their incidence increases during the last years, corresponding to the growth of air traffic and mainly to the increase of long duration without stop flight."[27] **c**, Chronology of various guidelines on HRT. The HRT was recommended consideration for elder women in the 1990s, but its effectiveness was questioned in the HERS trial (1998)[31]. Also, the risk was discovered in the WHI trial at the interim analysis (2002)[32]. It might be the effect of the publication on the HERS study (1998)[31] that the increase in thromboses was relatively small in 1998 despite the publication of two documents recommended in 1997. Panel a was reproduced from Clérel & Caillard. Syndrome thrombo-embolique de la station assise prolongée et vols de longue durée: l'expérience du Service Médical d'Urgence d'Aéroports De Paris. *Bull Acad Natl Med*. 1999; 183(5):985-997. discussion 997-1001. Figure 1. Copyright © 1999 Elsevier Masson SAS. All rights reserved. Académie Nationale de Médecine.

**Fig. S6.**

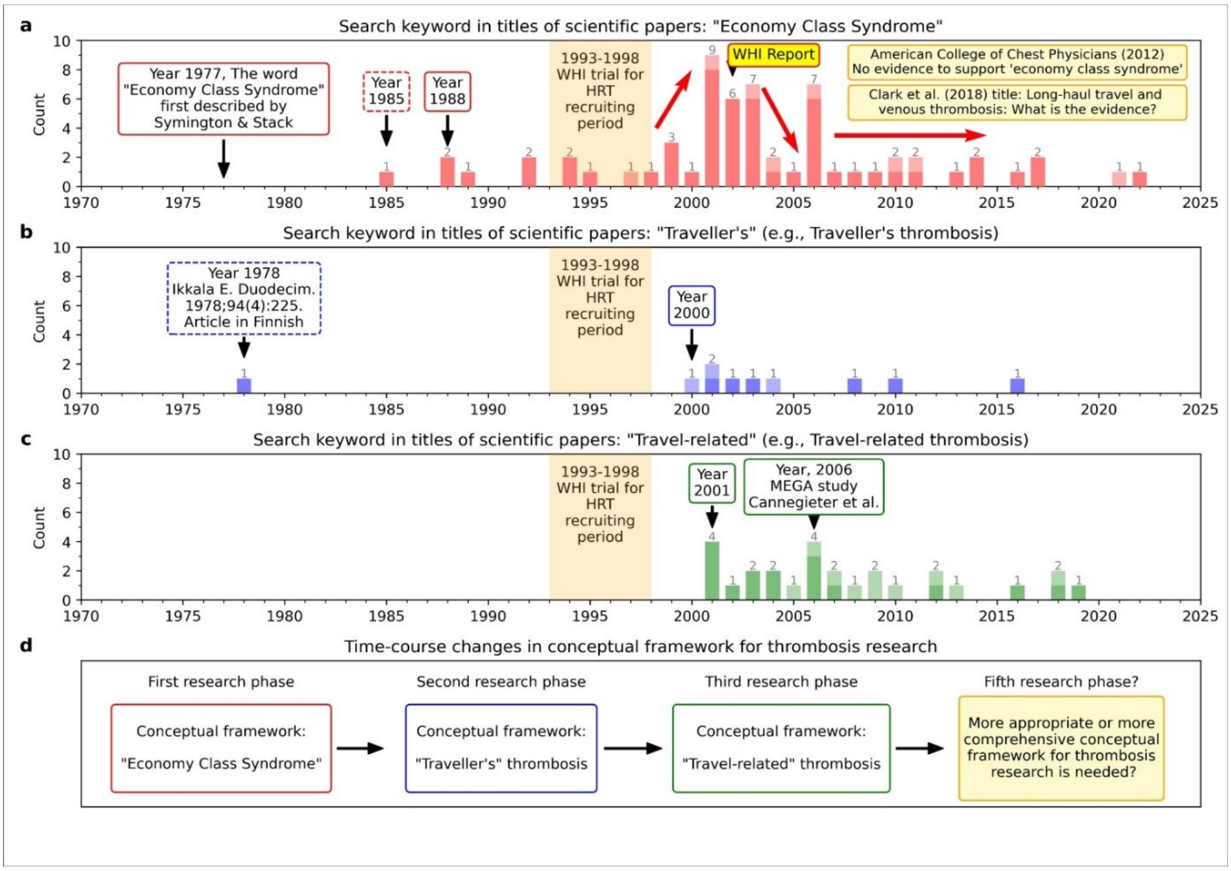


**Fig. S6. Visualized the change (trend) in the number of papers (trend analysis).**

**a**, Search results in the keyword “Traveler’s thrombosis.” **b**, Search results in the keyword “Traveler’s thrombosis” **c**, Search results with the keyword “Travel-related thrombosis”. **d**, Flow of the concept’s transition history. The bar graphs are based on the date of the electronic version. If PubMed displayed “Epub,” we choose the date of electronic version was opened, even if another publication day was described because the day of the electronic version is closer to the onset day. Light-colored bars indicate the review or meta-analysis articles.

**Fig. S7.**

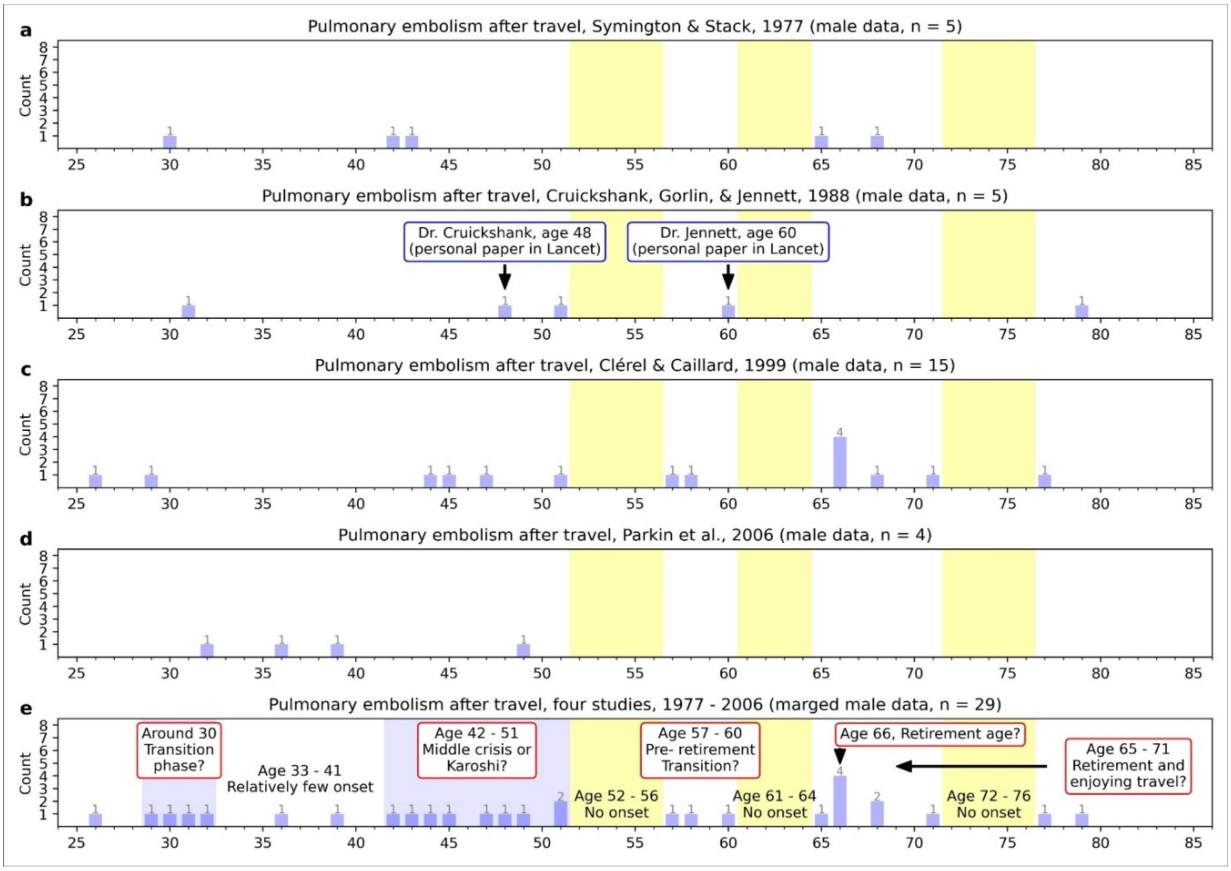


**Fig. S7. Displaying case data as a bar chart "by 1-year" after accumulating from several studies (male).**

**a**, Male data in Symington & Stack (1977)[33]. **b**, Male data in Cruickshank, Gorlin, & Jennett (1988)[34]. **c**, Male data in Clérel & Caillard (1999)[27]. **d**, Male data in Parkin et al (2006)[26]. **e**, Merged data from four studies[26,27,33,34]. Data on male cases extracted from articles on pulmonary embolism (PE) were published from the 1970s to the 2000s. In psychology, the transition period which adolescence to adulthood is around age 25. However, the study period is becoming longer with the increasing number of college students. It may shift to around the age of 30. In the data of Clérel & Caillard (1999)[27], four cases were recorded at age 66. Regarding the retirement age being extended to 65, this may be a case of a couple traveling to Paris after retirement.

**Fig. S8.**

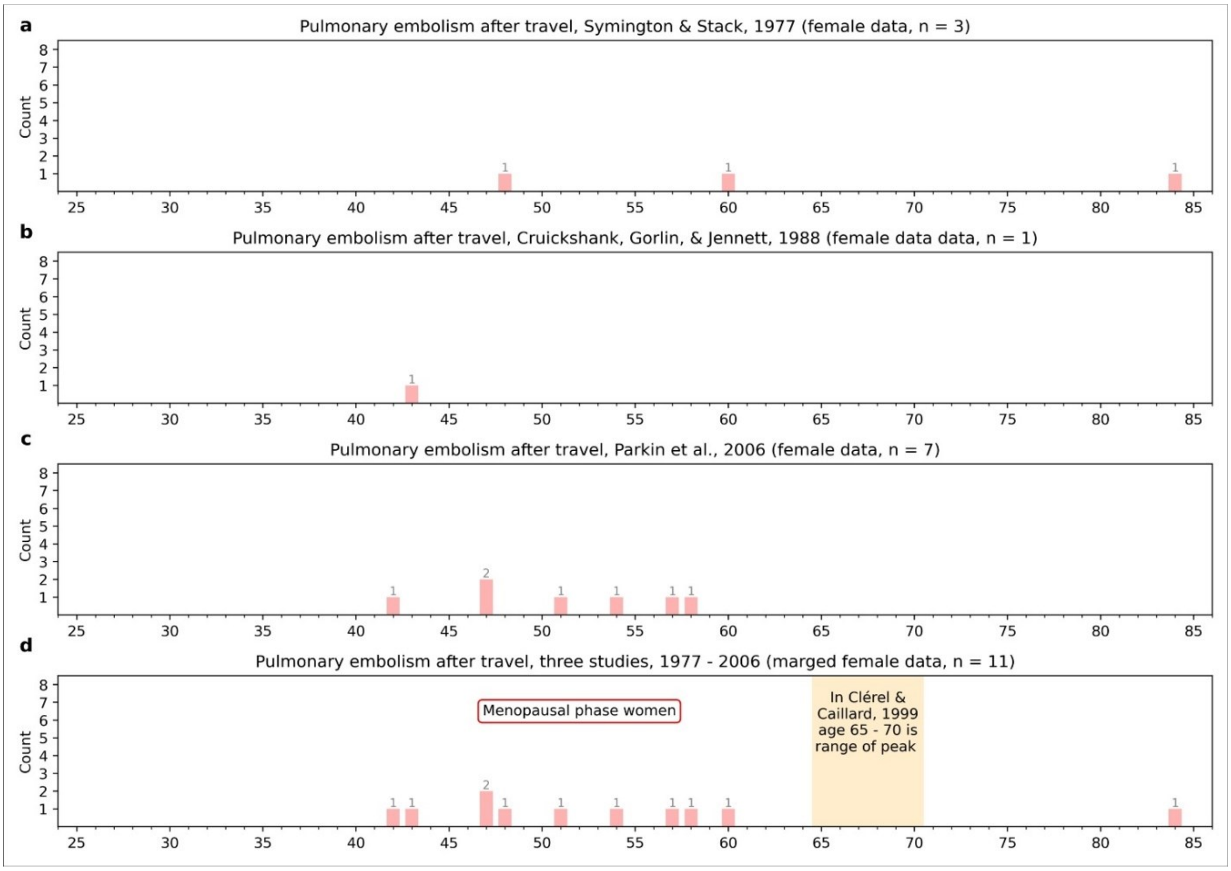


**Fig. S8. Displaying case data as a bar chart "by 1-year" after collecting from several studies (female).**

**a**, Female data in Symington & Stack (1977)[33]. **b**, Female data in Cruickshank, Gorlin, & Jennett (1988)[34]. **c**, Female data in Parkin et al (2006)[26]. **d**, Aggregated data from four studies. Data on female cases extracted from articles on pulmonary embolism (PE) were published from the 1970s to the 2000s.

**Fig. S9.**

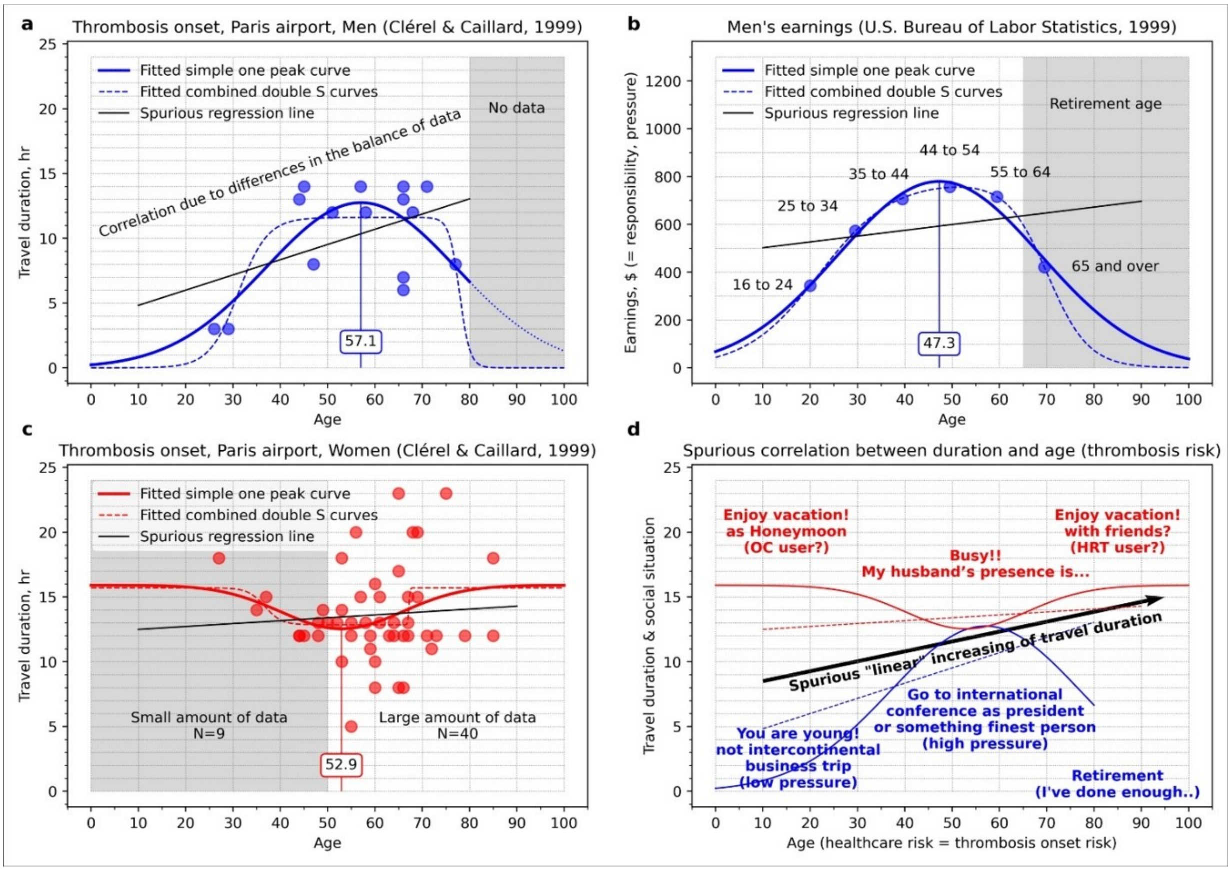


**Fig. S9. Relationship between age and travel duration in the data reported by Clérel & Caillard (1999).**

**a**, Scatterplot for the men's data included in the study reported by Clérel & Caillard (1999)[27]. Age on the horizontal axis and air travel time on the vertical axis. The peak position was 57.1 years old. **b**, Scatter plots of income in men in the United States (Bureau of Labor Statistics, U.S. Department of Labor, The Economics Daily, Men's earnings at peak at age 45-54 at https://www.bls.gov/opub/ted/1999/jul/wk3/art05.htm)[51]. Age is on the horizontal axis, and income is on the vertical axis. The peak position was 47.3 years old. **c**, Scatter plots for women's data in the study reported by Clérel & Caillard (1999) [27]. Age on the horizontal axis and air traveling time on the vertical axis. The position of the valley of the U-shaped curve was 52.9 years old. **d**, Illustration of the interpretation of the curves (inverted U-shape in men and U-shape in women) that are inverted each other. In all panels, the solid line fits the same curve as the normal distribution. The dotted line fits combined two S-shaped curves (curves represented by sigmoid functions). The difference between peak positions in an age (panels a and b) may be caused by the difference in data type. Air travel data may include a larger number of brain workers than income data due to the income data being for all men in the United States (e.g., peak is shifted to a later age for a more extended education period). In panel b, we have re-drawn the referenced material with reference to Section 105 of the U.S. Copyright Act and with confirmation that the referenced material is not copyrighted.

**Fig. S10.**

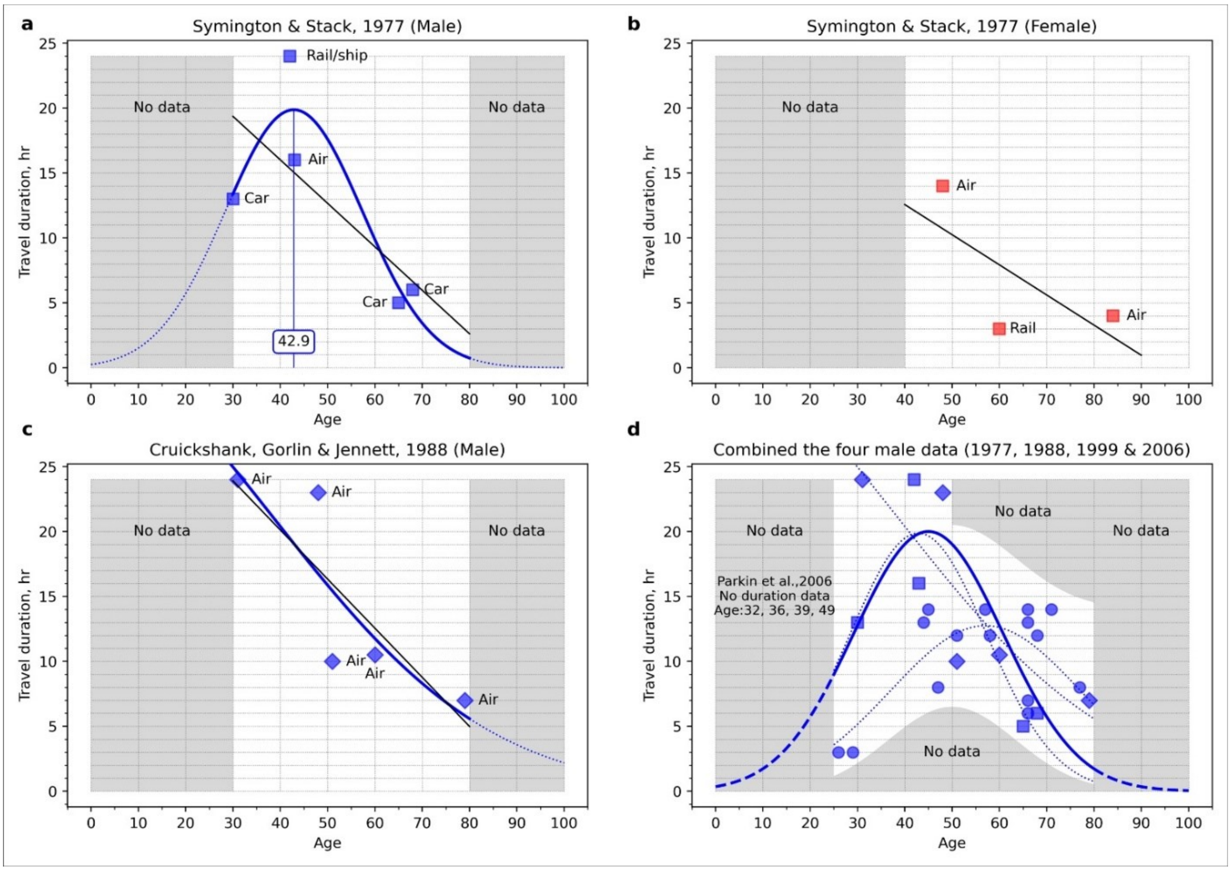


**Fig. S10. Relationship between age and duration in the data reported by four studies[26,27,33,34].** **a**, Extracting the data for male cases from Symington & Stack (1977)[33] and plotting them as age on the horizontal axis and travel time on the vertical axis. **b**, Extracting the data for female cases from Symington & Stack (1977)[33] and plotting them as age on the horizontal axis and travel time on the vertical axis. **c**, Extracting the data for male cases from Cruickshank, Gorlin, & Jennett (1988)[34] and plotting them as age on the horizontal axis and travel time on the vertical axis. **d**, Scatter plot of the data from four studies data. The data from Parkin et al (2006)[26] did not include travel time, but age information was described. The pattern in the scatterplot is not expected to change significantly. As for the data for women, we cannot make a judgment due to the small number of cases, but we suspect that the pattern may differ from that of Clérel & Caillard (1999)[27] for Paris airports **(c.f. Fig. S9, c & d)**.

**Fig. S11**

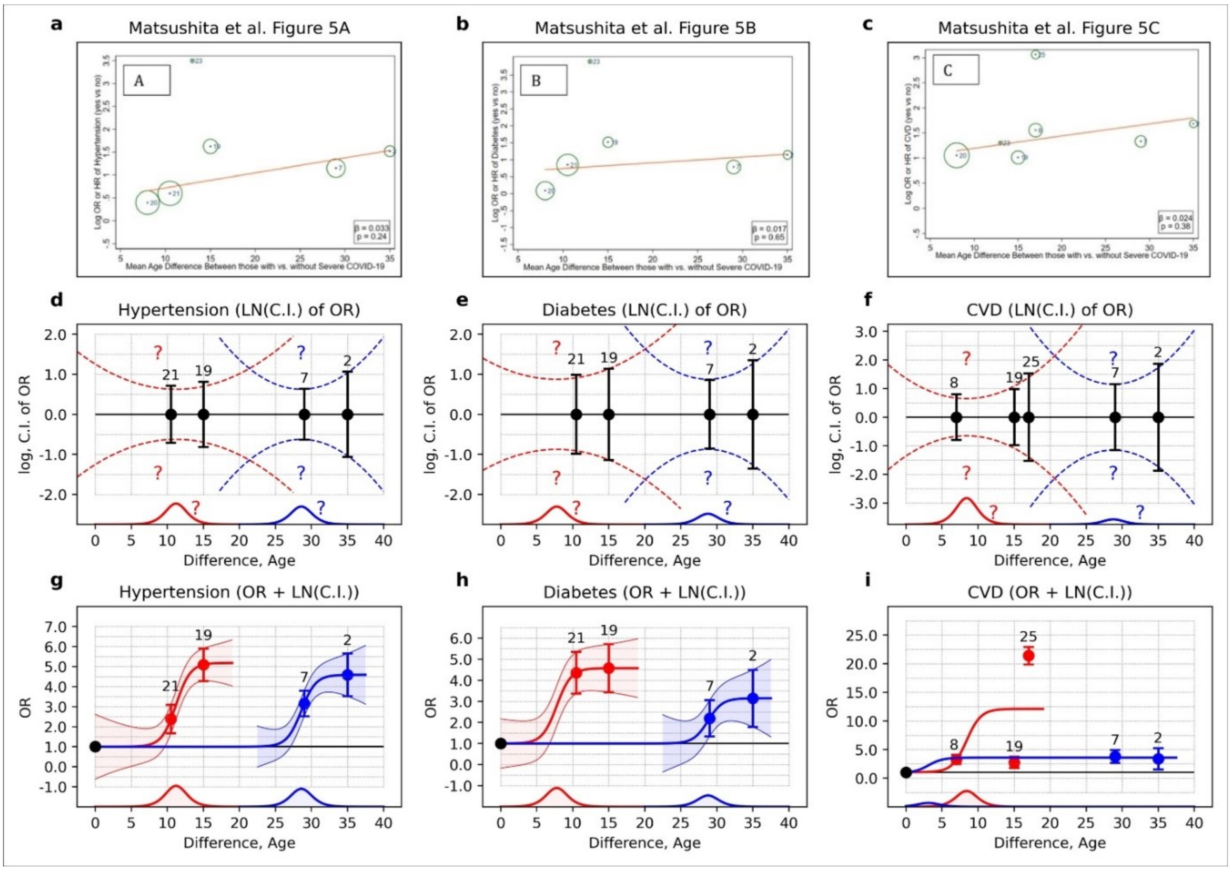


**Fig. S11. Hyperbolic shapes found in the figure reported by Matsushita et al[36].**

**a**, A figure reported by Matsushita et al shows the relationship between the severe and non-severe groups[36], which shows age difference on the horizontal axis and odds ratio (OR) or hazard ratio of hypertension on the vertical axis. To evaluate potential confounding for relative risk by age, Matsushita et al conducted meta-regression analyses based on the assumption that there was the possibility of confounding by age in the case that the study with a larger age difference has a higher relative risk[36]. **b**, Diabetes. **c**, Cardiovascular disease (CVD). **d-f**, Comparisons of error bars, which show 95% confidence interval (C.I.) s. It corresponds to the upper figure. **g-i**, Hyperbolic patterns were fitted to the OR and the 95% confidence limit of the OR. In the panel i, a hyperbolic shape could not be fitted due to the considerable data variation, likely due to the inconsistency of the term “CVD” (see **Supporting information: Method details**). The numbers marked at each point are the same as the numbers shown in the original figure. The sources of each data are shown in Methods. This figure was re-used from Matsushita et al. *Glob Heart*. 2020; 15(1):64. Figure 5. https://www.ncbi.nlm.nih.gov/labs/pmc/articles/PMC7546112/figure/F5/

Copyright © 2020 The Authors. Creative Commons Attribution 4.0 International License (CC-BY 4.0) https://creativecommons.org/licenses/by/4.0/

**Fig. S12.**

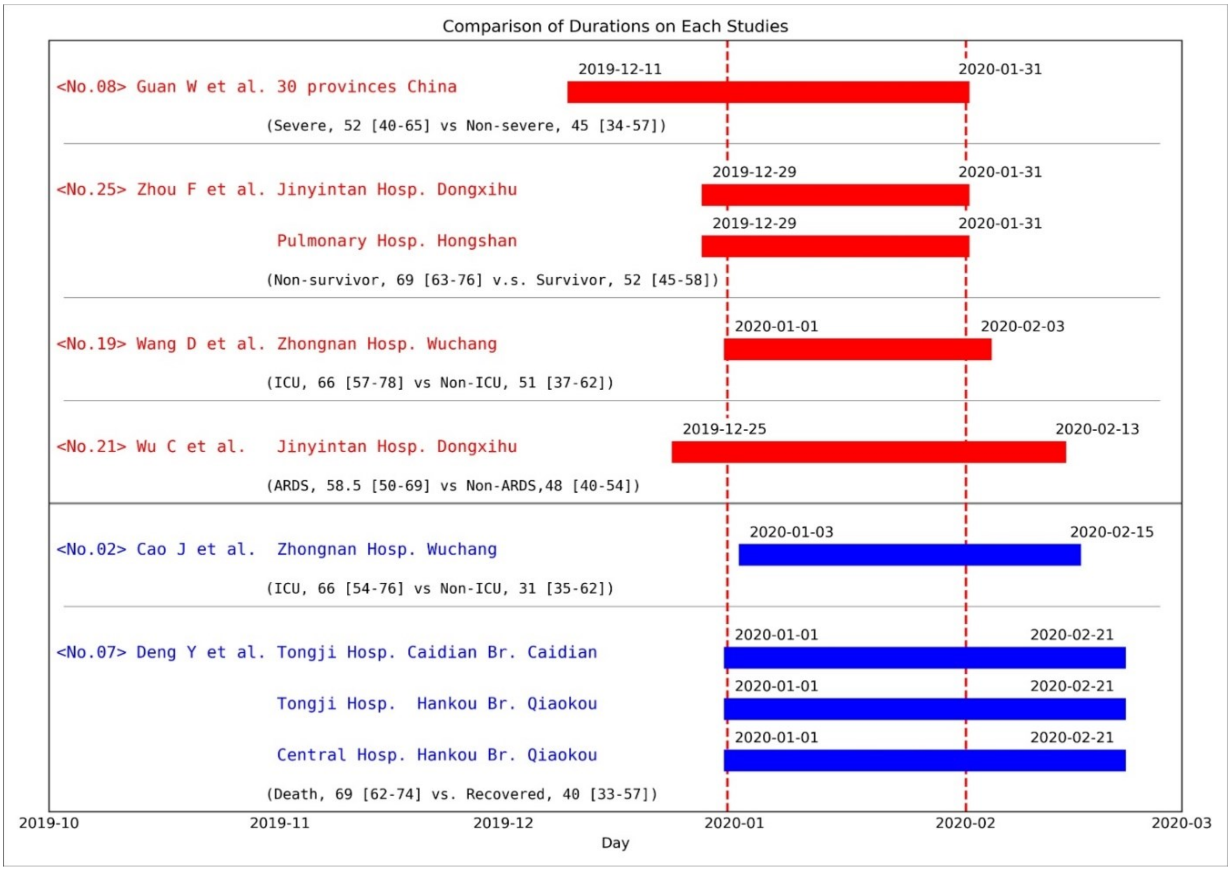


**Fig. S12. Data cut-off dates in each study cited by Matsushita et al[36].**

The data acquisition period in each study was displayed in Gantt chart format. The date display format is year-month-day. The correspondence between numbers and authors is as follows: No.2: Cao et al. (Cao, J. et al., *Intensive Care Med*. 2020;46(5):851-853.)[52], No.7: Deng et al (Deng, Y. et al., *Chin Med J (Engl)*. 2020;133(11):1261-1267.)[53], No.8: Guan et al (Guan, W.J. et al., *N Engl J Med*. 2020;382(18):1708-1720.)[54], No.19: Wang D. et al (Wang, D. et al., *JAMA*. 2020;323(11):1061-1069.)[55], No.20: Wang L. et al (Wang, L. et al., *J Infect*. 2020;80(6):639-645.)[56], No.21: Wu et al. (Wu, C. et al., *JAMA Intern Med*. 2020;180(7):934-943.)[57], No.25: Zhou et al (Zhou, F. et al., *Lancet*. 2020;395(10229):1054-1062.)[58]. Abbreviations: ARDS (Acute Respiratory Distress Syndrome), ICU (Intensive Care Unit). The number in parentheses means median age. The number in the bracket represents the standard deviation or interquartile range.

**Fig. S13.**

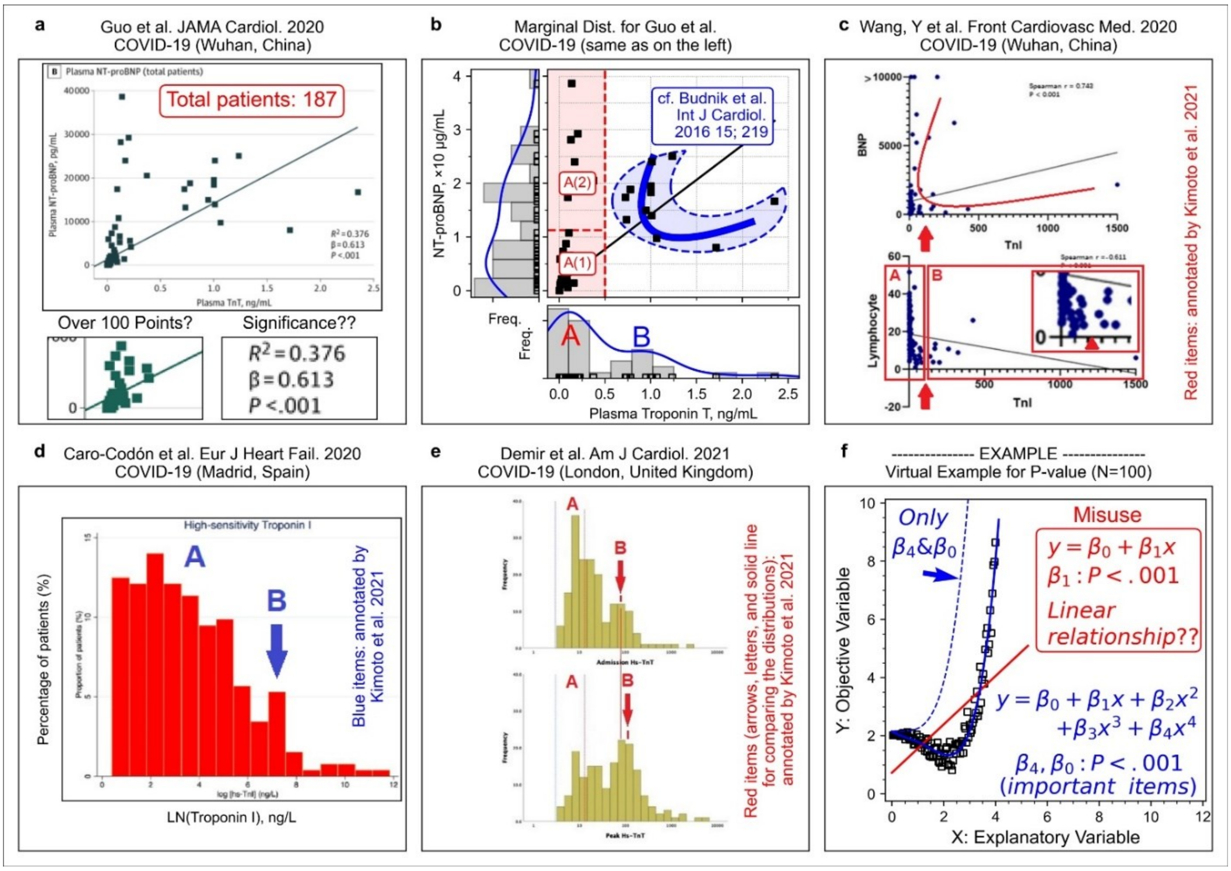


**Fig. S13. Bimodal distributions & tilted parabola in COVID-19 patients.**

**a**, Guo et al. *JAMA Cardiol*. 2020; 5(7):811-818. Figure 1B https://www.ncbi.nlm.nih.gov/labs/pmc/articles/PMC7101506/figure/hoi200026f1/ Copyright © 2020 Guo T et al. *JAMA Cardiology*. Creative Commons Attribution License (CC-BY). **b**, Marginal distribution of the scatter plot data. **c**, Wang et al. *Front Cardiovasc Med*. 2020; (7): 147. Figure 1 (upper, x-axis: troponin I, pg/mL; y-axis: BNP, pg/mL; lower, x-axis: troponin I, pg/mL; y-axis: lymphocyte, %)[39]. https://www.ncbi.nlm.nih.gov/labs/pmc/articles/PMC7477309/figure/F1/ Copyright © 2020 Wang, Zheng, Tong, Wang, Lv, Xi and Liu. CC BY License. **d**, Caro-Codón et al. *Eur J Heart Fail*. 2021; 23(3):456-464. Figure 1B (x-axis, LN (troponin I))[42]. https://www.ncbi.nlm.nih.gov/labs/pmc/articles/PMC8013330/figure/ejhf2095-fig-0001/ Copyright © 2021 European Society of Cardiology. All rights reserved. This Figure can be used for unrestricted research re-use and analysis in any form or by any means with acknowledgment of the original source as part of the COVID-19 public health emergency, for the duration of the emergency.

**e**, Demir et al. *Am J Cardiol*. 2021; 147:129-136. Figure 2 (upper: admission; lower: peak measurements; x-axis: troponin T, ng/L)[43]. https://www.ncbi.nlm.nih.gov/labs/pmc/articles/PMC7895690/figure/fig0002/ Copyright © 2021 Elsevier Inc. All rights reserved. This figure is granted for unrestricted research re-use and analyses in any form or by any means with acknowledgment of the original source by Elsevier for as long as the COVID-19 resource center remains active. **f**, Virtual example on regression analysis (**Supporting discussion 5: linear regression analysis**). In panel b (also c, d, and e), the histogram was bimodal (marked “A” and “B”). The crescent-shape pattern closely resembled the ST-segment elevation myocardial infarction group pattern that appeared in the study by Budnik et al[38].

**Fig. S14.**

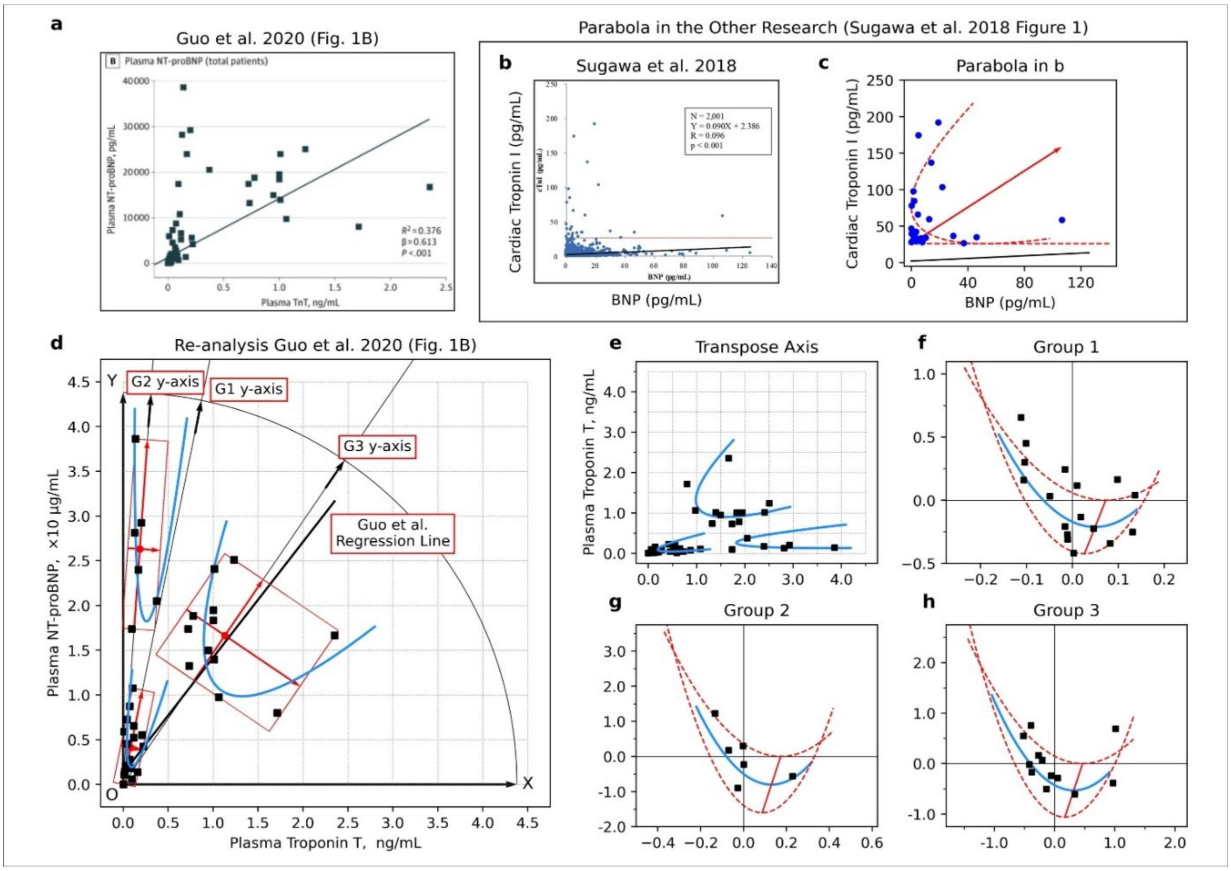


**Fig. S14. Three subgroup patterns appeared in a figure reported by Guo et al[37].**

**a**, A scatter plot showing the relationship between cardiac troponin T (TnT) and N-terminal pro-brain natriuretic peptide (NT-proBNP) in a patient with COVID-19[37]. **b**, Scatter plot to investigate the relationship between brain natriuretic peptide (BNP) and cardiac troponin I (cTnl) in healthy subjects reported by Sugawa et al[41]. **c**, The visible points that exceeded the value of 26.2 pg/mL (red line in panel b) were re-plotted with a parabola (not accurate regression analysis). **d**, Visible points in the figure reported by Guo et al[37] with parabolas. **e**, Transposed panel d for easy comparison. **f**, Group 1 in the small coordinate system (center of gravity as the origin of the coordinate system). **g**, Group 2 in the small coordinate system. **h**, Group 3 in the small coordinate system. In panel f-h, the upper curve is expressed by a quadratic function, in which a coefficient of the quadratic term equals a value of the coefficient of the quadratic term for the solid curve multiplied by 3/2. In the lower curve, the coefficient of the quadratic term of the solid curve multiplied by 2/3. Most data points located inside the crescent-shaped region enclosed by the parabolas, but the reason was unclear. Panel a was re-used from Guo T et al. *JAMA Cardiol*. 2020; 5(7):811-818. Figure 1B https://www.ncbi.nlm.nih.gov/labs/pmc/articles/PMC7101506/figure/hoi200026f1/ Copyright © 2020 Guo T et al. *JAMA Cardiology*. Creative Commons Attribution License (CC-BY). https://creativecommons.org/licenses/by/4.0/ Panel b was re-used from Sugawa et al. *Sci Rep*. 2018; 8(1):5120. Figure 1. https://www.ncbi.nlm.nih.gov/labs/pmc/articles/PMC5865159/figure/Fig1/ Copyright © 2018 The Authors. CC-BY 4.0 License

**Fig. S15.**

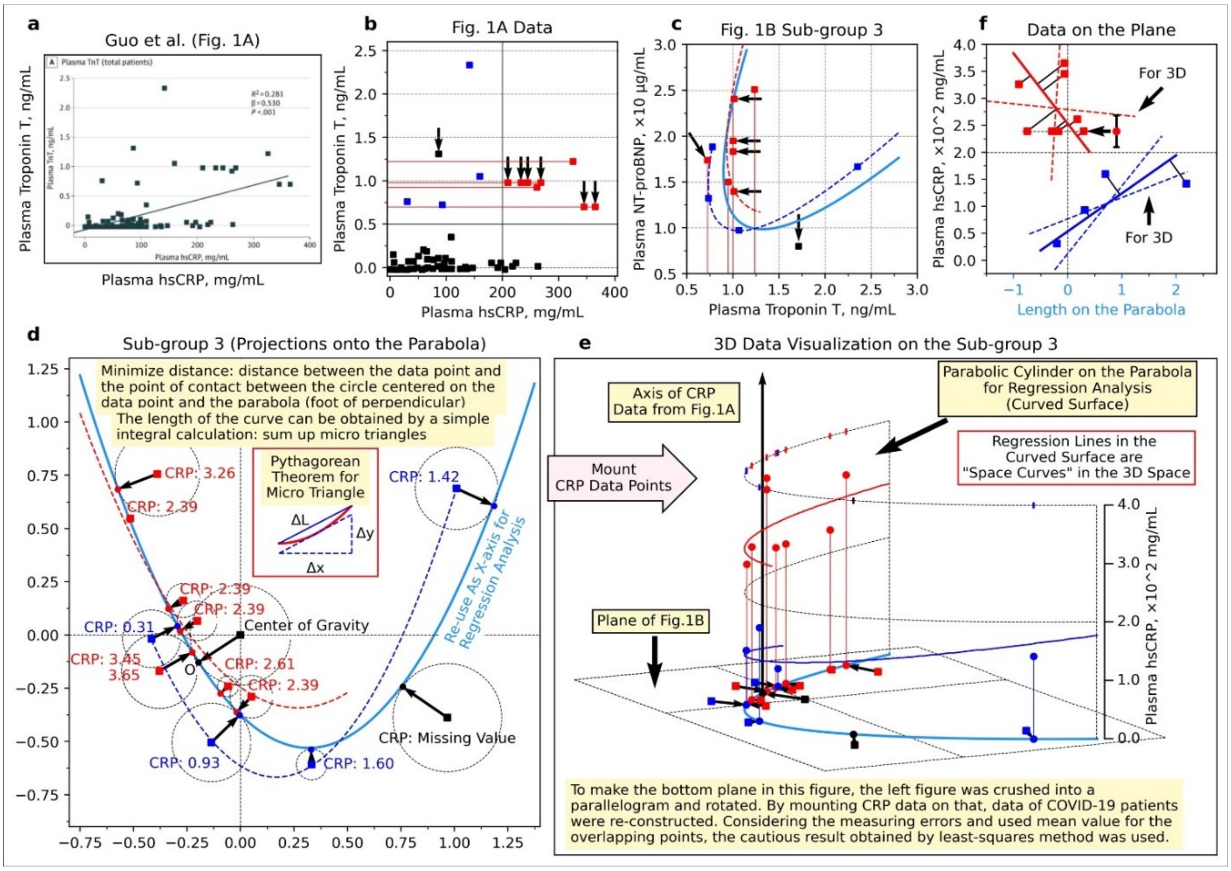


**Fig. S15. A three-dimensional plot reconstructed from the data reported by Guo et al[37].**

**a**, Scatter plot showing the relationship between high sensitive C-reactive protein (hsCRP) and troponin T (TnT)[37]. **b**, Re-drawn scatter plot with a vertical line around hsCRP = 200 mg/mL = 2.0 × 102 mg/mL. **c**, Enlarged subgroup 3 in panel d of **Fig. S14** (data exceeding hsCRP = 200 mg/mL are indicated by red, data not exceeding hsCRP = 200 mg/mL are indicated by blue). **d**, Enlarged subgroup 3 of panel d in **Fig. S14** with the foot of the perpendicular from each data point to the parabola. **e**, The hsCRP value of each patient placed on panel d (shown as a three-dimensional plot). **f**, The side surface of the parabolic cylinder. The point of TnT = 1.31 observed in panel b is not included in panel c, and the point of TnT = 1.71 in panel c is not included in panel b (inconsistent). In panel e, the projection of the data points onto the parabola was used as new points. In panels e and f, some points of CRP value could not be determined because of overlapping, so those points were replaced with the average value (the points indicated by the left-pointing arrows and the error bar, which are the average values and the range of values, respectively). Panel a was re-used from Guo et al. *JAMA Cardiol*. 2020; 5(7):811-818. Figure 1. https://www.ncbi.nlm.nih.gov/labs/pmc/articles/PMC7101506/figure/hoi200026f1/ Copyright © 2020 Guo T et al. *JAMA Cardiology*. Creative Commons Attribution License (CC-BY). https://creativecommons.org/licenses/by/4.0/

### 2. Supporting tables & table legends

#### 2.1. Supporting Table 1: data review results

To confirm the accuracy of the values, we re-calculated the odds ratio (OR) and confidence interval of the OR. In the data reported by Kuipers et al, we re-calculated the Incidence Ratio (IR). However, Kuipers et al[5] did not describe a method for calculating the confidence interval of the IR (e.g., exact method or not). So we could not calculate the confidence interval. The OR, confidence interval of the OR, and IR were re-calculated with Microsoft Excel®. The values were rounded by a rounding function such as “ROUND()”. The result is shown in following **Table S1**.

**Table S1. data review results.**

| Ref. | hour | a | b | c | d | Re-calculated OR or IR | | | | | Described OR or IR | | | | | Difference | | | | |
|---|---|---|---|---|---|---|---|---|---|---|---|---|---|---|---|---|---|---|---|---|
| | | | | | | OR | | CIL | CIU | | OR | | CIU | CIL | | | | | | |
| C | 4-8 | 36 | 18 | 1,673 | 1,724 | 2.00 | ( | 1.14 | 3.52 | ) | 2.0 | ( | - | - | ) | 0.00 | ( | - | - | ) |
| C | 8-12 | 33 | 18 | 1,673 | 1,724 | 1.83 | ( | 1.03 | 3.25 | ) | 1.8 | ( | - | - | ) | 0.03 | ( | - | - | ) |
| C | >12 | 25 | 9 | 1,673 | 1,724 | 2.78 | ( | 1.30 | 5.96 | ) | 2.8 | ( | - | - | ) | -0.02 | ( | - | - | ) |
| K | 0–4 | 2 | 4,267 | 29 | 27,772 | 0.47 | ( | - | - | ) | 0.5 | ( | 0.0 | 1.4 | ) | -0.03 | ( | - | - | ) |
| K | 4–8 | 5 | 2,180 | 29 | 27,772 | 2.29 | ( | - | - | ) | 2.3 | ( | 0.7 | 4.8 | ) | -0.01 | ( | - | - | ) |
| K | 8-12 | 6 | 2,676 | 29 | 27,772 | 2.24 | ( | - | - | ) | 2.2 | ( | 0.8 | 4.4 | ) | 0.04 | ( | - | - | ) |
| K | 12-16 | 7 | 1,344 | 29 | 27,772 | 5.21 | ( | - | - | ) | 5.2 | ( | 2.0 | 9.9 | ) | 0.01 | ( | - | - | ) |
| K | >16 | 4 | 672 | 29 | 27,772 | 5.95 | ( | - | - | ) | 5.9 | ( | 1.5 | 13.4 | ) | 0.05 | ( | - | - | ) |
| M | Short & Long | 31 | 16 | 179 | 194 | 2.10 | ( | 1.11 | 3.97 | ) | 2.1 | ( | 1.1 | 4.0 | ) | 0.00 | ( | 0.01 | -0.03 | ) |
| M | Short: 2 | 20 | 12 | 179 | 194 | 1.81 | ( | 0.86 | 3.81 | ) | - | ( | - | - | ) | - | ( | - | - | ) |
| M | Long 13.5 | 11 | 4 | 179 | 194 | 2.98 | ( | 0.93 | 9.53 | ) | 3.0 | ( | 0.9 | 9.5 | ) | -0.02 | ( | 0.03 | 0.03 | ) |
| P | >3 | 5 | 9 | 83 | 325 | 2.18 | ( | 0.71 | 6.68 | ) | 2.2 | ( | 0.7 | 6.8 | ) | -0.02 | ( | 0.01 | -0.12 | ) |
| P | 3-8 | 0 | 5 | 83 | 325 | | ( | | | ) | - | ( | - | - | ) | - | ( | - | - | ) |
| P | >8 hr. | 5 | 4 | 83 | 325 | **4.89** | ( | **1.28** | **18.61** | ) | **6.0** | ( | **1.4** | **25.4** | ) | **-1.11** | ( | **-0.12** | **-6.79** | ) |

Abbreviations: C: Cannegieter et al, K: Kuipers et al, M: Martinelli et al, P: Parkin et al, OR: Odds Ratio, IR: Incidence Ratio, CIL: end of confidence interval (lower limit), CIU; end of confidence interval (upper limit).

### 2.2. Supporting Table 2: verification of a calculated value by Parkin et al.

In the value reported by Parkin et al[26], there was a discrepancy between the OR and the described OR calculated by us. Although Parkin et al might have used unique specifications of the analysis software (SAS), it was suspected that cells in the cross table were mistaken (*e.g.*, in the table of Table S2, a, the cell for control and the cell for total was mistaken). However, the error bar's length was relatively small since the total number of cases was notably smaller than other studies. So, qualitatively, it could be used to group data to find hyperbolic patterns.

**Table S2. Verification of a calculated value by Parkin et al.**

| Flight details | Cases | Controls | Row marginal total |
|---|---|---|---|
| **No long-distance flight** | 83 | **325** | 83+325=**408** |
| **Air travel 8 > hours** | 5 | 4 | 5 + 4 = 9 |
| **Column marginal total** | 83 + 5 = 88 | 325 + 4 = 329 | 83 + 5 = 88 |

The data in each cell were derived from Parkin et al.

(Parkin et al. *Thromb Haemost* 2006; 95: 807-814, Table 5).

$$Odds\ ratio = \frac{\mathbf{325} \times 5}{83 \times 4} \cong 4.89$$

$$Calculation\ using\ a\ value\ of\ row\ marginal\ total = \frac{\mathbf{408} \times 5}{83 \times 4} \cong 6.0$$

$$= described\ value\ by\ Parkin\ et\ al.$$

### 2.3. Supporting Table 3: Changing female percentage from four studies reported in the 1970s to 2000s.

**Table S3. Changing female percentage from four studies reported in the 1970s to 2000s.**

| Author | Year | Total N | Male | | Female | | Larger one |
|---|---|---|---|---|---|---|---|
| | | | Male N | % | Female N | % | |
| Symington & Stack[33] | 1977 | n=8 | n=5 | 62.5% | n=3 | 37.5% | Male |
| Cruickshank et al[34] | 1988 | n=6 | n=5 | 83.3% | n=1 | 16.7% | Male |
| Clérel & Caillard[27] | 1999 | n=64 | n=15 | 23.4% | n=49 | 76.6% | Female |
| Parkin et al[26] | 2006 | n=11 | n=4 | 36.4% | n=7 | 63.6% | Female |

### 3. Supporting information: Method details

#### 3.1. Deriving the formula for regression analysis

##### 1) Mathematical considerations

Deriving the exact formula directly was difficult. So, firstly we focused only on the U-shaped curve. The length of the confidence interval for the odds ratio is inverse proportional-like to the size of the potential patient population, which onset thrombosis by exposure to risk factors **(see Fig. S2)**. Although, there are multiple possibilities (e.g., 1/N, $1/N^2$, or 1/sqrt(N)), regarding the form of equations (1) and (2) below, we intuitively predicted that it would be approximately equal to the inverse of the root (equation (3)).

$$Log_e(95\%CI) = Log_e\,(OR) \pm 1.96\sqrt{\frac{1}{a}+\frac{1}{b}+\frac{1}{c}+\frac{1}{d}} \quad (1)$$

$$\sqrt{\frac{1}{a}+\frac{1}{b}+\frac{1}{c}+\frac{1}{d}} = \frac{\sqrt{bcd+acd+abd+abc}}{\sqrt{abcd}} = \frac{complex\ combination\ of\ numbers}{\sqrt{abcd}} \quad (2)$$

$$Log_e(95\%CI) = Log_e\,(OR) \pm 1.96\sqrt{\frac{1}{a}+\frac{1}{b}+\frac{1}{c}+\frac{1}{d}} \cong Log_e(OR) \pm 1.96 \times \frac{1}{\sqrt{N}} \quad (3)$$

To be precise, the ability of patient recruitment, in other words, the "sample size in each study," affects the confidence interval length.

It is similar to Hooke's law on spring[59], which states that the pull-back force of spring is proportional to the length of the pull. In this case, the power also depends on the spring type (e.g., thick spring or thin spring). Thus, the formula is as follows, with spring constant (k) depending on the kind of spring (equation (4)). Similarly, equation (3) express equation (5) precisely.

$$Force = k \times Distance \quad (4)$$

$$Log_e(95\%CI) = Log_e\,(OR) \pm 1.96\sqrt{\frac{1}{a}+\frac{1}{b}+\frac{1}{c}+\frac{1}{d}} \cong Log_e(OR) \pm 1.96 \times \frac{k}{\sqrt{N}} \quad (5)$$

However, we speculated that the effect of the differences in sample size could not be a practical problem for two reasons.

The first reason is that stratified analysis, often performed in clinical studies, can be a force in reducing the differences. The second reason is that two types of boundary conditions in the hyperbolic shape fitting, which are the "S-curve" and the "error bar length," reduce the degree of freedom in searching and increase the possibility of finding the desired shape.

### 2) Numerical Experiment

Usually, a sigmoid function is used for curve fitting to a dose-response relationship and outline of the cumulative distribution. Also, the curve expressing the end of a confidence interval is a sum of the S-shaped and U-shaped curves **(see Fig. 1)**.

To decide on a formula to express a U-shaped curve, we performed a numerical experiment. The result indicated that a parabola could approximate a U-shaped curve **(Fig. 1, c & d).** We constructed three mathematical formulas for fitting hyperbolic patterns to data **(see Fig. 1).**

### 3) Identifying the formula

Considering the numerical experiment, we decided on the equations for the S-shaped curve, the upper end of the confidence limit, and the lower end of the confidence limit (equation (1), (2), and (3)). Note that in the below equations, the coefficient L is generally set as 1. So, we used this equation under the condition of L=1 unless there is some reason.

$$y = \left\{\frac{K_1}{1+e^{L(x-M)}} + K_2\right\} \quad (6)$$

$$y = \left\{\frac{K_1}{1+e^{L(x-M)}} + K_2\right\} + (ax^2 + bx + c) \quad (7)$$

$$y = \left\{\frac{K_1}{1+e^{L(x-M)}} + K_2\right\} - (ax^2 + bx + c) \quad (8)$$

#### 3.2. Tools used in this research.

**1) Numerical experiment**

To generate binomial distribution data, we used the “BINOM.DIST” function, which is a function for calculating the probability of binomial distribution in a kind of spreadsheet software, Microsoft Excel® (Microsoft Corporation, Redmond, Washington, US).

**2) Regression analysis**

Regression analysis was performed using Python (Python Software Foundation, Delaware, USA https://www.python.org/psf/records/incorporation/). At this time, Python's functional modules NumPy (NumFOCUS sponsored open-source project, https://numpy.org/), Pandas (NumFOCUS sponsored open-source project, https://pandas.pydata.org/), SciPy (NumFOCUS sponsored open-source project, https://www.scipy.org/) and Matplotlib (NumFOCUS sponsored open-source project, https://matplotlib.org/) were also used.

Additionally, a function as “Chart option” of Microsoft Excel®, which was shown in the "Trendline Options" section contained in “Format Trendline,” was used.

**3) Reading values in the figures**

Reading values from the published figures were performed using the public domain software ImageJ (https://imagej.net/Welcome).

**4) Other tools**

When symbolic formula manipulation was required, formula manipulation software wxMaxima (Project Maxima maintained by 27 volunteers, https://maxima.sourceforge.io/) was used.

#### 3.3. Analysis 1A (dataset: Chandra et al)[10]

**1) Data review to validate eligibility for regression analysis**

a. The process of data review in this analysis

As the first step, we performed data mapping. In the figure of meta-regression analysis reported by Chandra et al[10], it was not described what the data points correspond to the four original papers (Martinelli et al, 2003; Parkin et al, 2006; Cannegieter et al, 2006 and Kuipers et al, 2007)[1,3,5,26]. So, we measured the positions of each point and compared them with the original descriptions in the papers.

In the second step, we performed a data review, which was an examination of the accuracy of cited values, and appropriately from the viewpoint of biomedicine. Our re-calculation confirmed the odds ratio (OR), confidence interval of the OR, and adjusted OR. In examining the values, we did not confirm Chandra et al[10] and the four authors[1,3,5,26] because Chandra et al described that each author did not respond to the inquiry[10]. As a final step, we performed regression analysis using the eligible data for using regression analysis.

**b. Data review**

In Parkin et al[26], there was a discrepancy between the OR and the described OR calculated by us. Also, it was suspected that cells in the cross table were mistaken (*e.g.*, in the table of Fig. 1a, the cell for control and the cell for total were mistaken). However, the error bar's length was relatively small since the total number of cases was notably smaller than other studies. So, qualitatively, it could be used to group data to find hyperbolic patterns.

**2) Regression Analysis**

For the data judged as eligible, hyperbolic patterns were visually searched, and each data point was grouped into two groups. Then, a non-linear regression analysis using the formulas above was performed. In the fitting of the U-shaped curve, since there were many unknown coefficients for the number of data (there are seven unknowns, $K_1$, $K_2$, M, a, b, and c. in the equations (7) and (8)), the

S-curve was fitted first, and the remaining unknown coefficients (a, b, c) were fitted to the residuals of the S-curve fitting (see equation (7) and (8)).

**3) Additional analysis**

As described above, the cyclic pattern in the data reported by Cannegieter et al[3] was observed, and we performed additional analysis. To conduct an appropriate non-linear regression analysis, we made an equation by combining two types of equations. The exponential decay equation was usually used to express radioactive decay in physics and clearance in medicine. The other was a trigonometric function (sine function) to express waveforms. The equation is shown in as below equation (9). Also, this scientific model (model formula) was used to estimate the ratio of patients by integral calculation.

$$y = N_1 e^{-\lambda_1 (t-b)} + N_2 e^{-\lambda_2 (t-b)} \operatorname{Asin}\{B(t-b)\} \quad (9)$$

The data reported as a bar graph was weekly data **(see Fig. 4)**. So, the week was converted into the number of days before the analysis, such as; the day getting off the vehicle was set as days 0, the first week was set as days 4, the second week was set as days 4 + 7, and the third week was set as days 4 + 7 × two, and the Nth week was set as days 4 + 7 × N.

Supplementary, according to the original description by Cannegieter et al[3], 68 patients developed thrombosis in the first week, and “233” patients developed thrombosis within eight weeks after traveling. However, there was a slight discrepancy in the values read from the bar graph. The value in our measurement within eight weeks after traveling was “234”, but the effect of only one patient was allowed to be regarded as small. Cannegieter et al described that there were missing values [3]. The mismatch between our measurement and description might be related to the described explanation.

### 3.4. Analysis 1B (dataset: Philbrick et al) [7]

#### 1) Dataset search and data review

a. Dataset search

To validate the result of analysis 1A, we searched another dataset from a meta-analysis or systematic review of the traveler's thrombosis. To conduct this search, we used PubMed® setting the following search keywords: "economy class syndrome [Title] " OR "traveler's [Title] AND thrombosis [Title]" OR "traveler's [Title] AND thromboembolism [Title]" OR "flight [Title] AND thrombosis [Title]" OR "flight [Title] AND thromboembolism [Title]" OR "flight-related [Title] AND thrombosis [Title]" OR "flight-related [Title] AND thromboembolism [Title]" OR "travel [Title] AND thrombosis [Title]" OR "travel [Title] AND thromboembolism [Title]" OR "travel-related [Title] AND thrombosis [Title]" OR "travel-related [Title] AND thromboembolism [Title]" (Filters: Meta-Analysis, Systematic Review).

As a result, we obtained the eight articles (da Silva LF et al. *J Vasc Bras.* 2021 10;20:e20200164; Benhaberou-Brun *Perspect Infirm.* 2010 7(3):16-7; Chandra et al. *Ann Intern Med.* 2009 151(3):180-90; Kuipers et al. *J Intern Med.* 2007 262(6):615-34; Philbrick et al. *J Gen Intern Med.* 2007 22(1):107-14; Hsieh et al. *J Adv Nurs.* 2005 51(1):83-98; Ansari et al. *J Travel Med.* 2005;12(3):142-54; Adi et al. *BMC Cardiovasc Disord.* 2004 19;4:7).

Subsequently, we selected articles containing available abstracts on PubMed® online, confirming the contents. As a candidate for our analysis, we chose a systematic review reported by Philbrick et al. [7]. The study was taken up by the ACP Journal Club of the American College of Physicians [8]. So, it seemed to be a highly reputed study. Therefore, we regarded that the dataset contained in the research was suitable for validation.

Also, the research contained two lists of tables, one of which was a cohort studies dataset, and the other was case-control studies. However, the case-control studies had many different exposure factors. So, we decided to use only the cohort studies dataset.

b. Data review

In the data review process, we reviewed the table containing ten cohort studies[27,28,60-67] and found seven eligible cohort studies[27,60-63,65,66] for regression analysis. In Gajic et al and Kelman et al, only distances were described [28,67], and Hughes et al reported duration data for not per one flight (e.g., mean 39.4 h)[64], so time data for regression analysis was unavailable.

Additionally, although Philbrick et al described that incidence per million was 0.5 on the result reported by Clérel & Caillard[27], Clérel & Caillard mentioned, "According to the number of passengers landing in the Aeroports de Paris, the incidence during 1998 is 0.5 per million passengers"[27].

### 2) Regression Analysis

For the seven studies, data stratified by Pulmonary Embolism (PE) and Deep Vein Thrombosis (DVT), regression analysis was performed using an S-shaped curve formula (see equation (1)). In the case of curve-fitting on DVT data, we canceled the setting of coefficient L=1 to increase the degree of freedom of the S-curve **(Fig. 1, b)**. To show the error bar in the figure, we did not use the values of confidence limits described in the report by Philbrick et al[7], but values were re-calculated from the number of cases using Wilson's method.

In the seven studies, not OR or relative risk (RR), only the data indicating the incidence rate of thrombosis was available. So, the hyperbolic pattern did not appear in the figure theoretically, and we performed only the S-shaped curve fitting. This mechanism is explanted from the following calculation on a confidence interval of a ratio.

The formula for a 95% confidence limit of a ratio using binomial approximation is expressed by the following formula: P is a ratio, and N is the number of trials.

$$P - 1.96\frac{\sqrt{P(1-P)}}{\sqrt{N}} \leq P \leq P + 1.96\frac{\sqrt{P(1-P)}}{\sqrt{N}} \tag{10}$$

In the above equation, the fraction's numerator is not a constant value. It does not depend on only the N, which is associated with a data point's position in a population **(see Fig. S2).**

In this regression analysis, converting time categories to time points was necessary, so we performed this in three directions. The first one was taking the midpoint if the category was not the end of a category sequence (e.g., 10-15 h could be converted to 12.5 h). The second one took the midpoint between the time point of 0 and the lower limit of the category if the category was the lower end of a category sequence (e.g., <3 h could be converted to 1.5 h). The third one was taking the sum of the value of the upper limit and the value of the midpoint between the time point of 0 and the lower limit of the category sequence if the category was the upper side of a category sequence (e.g., > 12 h could be converted to 12 h + 1.5 h =13.5 h). Details of conversions are shown below (the original time category is shown in brackets).

Belcaro et al [10-15 h]: 12.5 h (Belcaro, G. et al., *Angiology.* 2001;52(6):369-74.)[60]; Clérel et al [12.7 h]: 12.7 h (Clérel, M., & Caillard, G., *Bull Acad Natl Med.* 1999;183(5):985-97.)[27]; Jacobson et al. [11 h]: 11 h; Lapostolle et al [<3 h, 3-6 h, 6-9 h, 9-12 h,> 12 h]: 1.5 h, 4.5 h, 7.5 h, 10.5 h, 13.5 h (12 + 1.5 = 13.5 h) (Jacobson, B.F. et al., *S Afr Med J.* 2003;93(7):522-8.) [63]; Pérez-Rodríguez et al [<6 h, 6-8 h,> 8 h]: 3 h, 7 h, 11 h (8 + 3 = 11 h) (Pérez-Rodríguez, E. et al., *Arch Intern Med.* 2003;163(22):2766-70.)[65]; Schwarz et al 2002 [> 8 h]: 12 h (midpoint of 0-8 h is 4 h and 8 + 4 = 12 hours) (Schwarz, T. et al., *Blood Coagul Fibrinolysis.* 2002;13(8):755-7.)[62]; Schwarz et al 2003 [> 8 h]: 12 h (midpoint of 0-8 h is 4 h and 8 + 4 = 12 hours) (Schwarz, T. et al., *Arch Intern Med.* 2003 2003;163(22):2759-64.)[66].

#### 3) Additional analysis

a. Regression analysis (data: Kelman et al[28])

In the review process, a cyclic pattern was observed. So, we worked on regression analysis. Considering that onset of thrombosis tends to increase again, an equation upward-sloping curve was added to equation (9). The equation is the following (11).

$$y = N_1 e^{-\lambda_1 (t-b)} + N_2 e^{-\lambda_2 (t-b)} \mathrm{A}\sin\{B(t-b)\} + (ax^2 + bx + c) \qquad (11)$$

b. Analysis by using correlogram (data: Clérel & Caillard[27])

We considered using the “correlogram” in this study because it was more practical than observing the original data's fluctuation. Periodic fluctuation patterns may be unclear when looking at the original data alone, but potential patterns can be obtained using a correlogram, a data visualization method for analyzing time-series data. Also, as the correlation coefficient plotted on the correlogram, we decided to use Spearman's rank correlation coefficient instead of Pearson's product-moment correlation coefficient, which is easily affected by outliers. Also, we performed a non-linear regression analysis using a mathematical formula (12) that includes two sine functions.

$$y = n_1 \sin\{a_1(x - b_1)\} + n_2 \sin\{a_2(x - b_2)\} \quad (12)$$

In correlogram creation, firstly, a combination of data (data X1, data X1) was created by arranging the original time series data (data X1), and a new combination (data X1, data X1') was created by shifting one of them. Secondary, the correlation coefficient (also called the auto-correlation coefficient) between the original time-series data (data X1) and the sifted time-series data (data X1'), except at the ends of two types of time-series data where some correspondence could not be formed. By repeating shifting the time string data and calculating the correlation coefficient, the locus of the correlation coefficient becomes the shape of waves. Firstly (original waves of time strings are overlapped), the correlation coefficient is 1, and the value of the correlation coefficient gradually decreases. Finally (the wave is inverted), the correlation coefficient is -1.

#### 3.5. Analysis 1B-related additional analysis

##### 1) Literature search

We searched the literature for trend analysis. To conduct this search, we used PubMed® setting the following search keywords: “economy class syndrome,” “traveler's thrombosis,” and “travel-related thrombosis.”

a. Details of the keyword (“economy class syndrome”)

Search: (economy[Title]) AND (class[Title]) AND (syndrome[Title])

b. Details of the keyword (“traveler's thrombosis”)

Search: ((traveller's[Title]) AND (thrombosis[Title])) OR ((traveller's[Title]) AND (thromboembolism[Title])) OR ((traveller's[Title]) AND (pulmonary[Title]) AND (embolism[Title])) OR ((traveller's[Title]) AND (deep[Title]) AND (vein[Title]) AND (thrombosis[Title])) OR ((traveller's[Title]) AND (venous[Title]) AND (thrombosis[Title])) OR ((traveller's[Title]) AND (venous[Title]) AND (thromboembolism[Title]))

c. Details of the keyword (“travel-related thrombosis”)

Search: ((travel-related[Title]) AND (thrombosis[Title])) OR ((travel-related[Title]) AND (thromboembolism[Title])) OR ((travel-related[Title]) AND (pulmonary[Title]) AND (embolism[Title])) OR ((travel-related[Title]) AND (deep[Title]) AND (vein[Title]) AND (thrombosis[Title])) OR ((travel-related[Title]) AND (venous[Title]) AND (thrombosis[Title])) OR ((travel-related[Title]) AND (venous[Title]) AND (thromboembolism[Title]))

2) Confirming citation history

To confirm citation history, we checked the “Citations & Impact” page on the Europe PMC website (https://europepmc.org/).

#### 3) Additional analysis

a. Data visualization

We displayed patient data as a bar chart "by one year" after collecting from several studies. Also, we made a scatter plot as age on the horizontal axis and travel time on the vertical axis.

b. Regression analysis

We performed regression analysis using the same curve as the normal distribution. In women, we use inversed curve. Also, we fitted combined two S-shaped curves represented by sigmoid functions.

$$y = Le^{-a(x-d)^2} \tag{13}$$

$$y = L_2 - L_1 exp(-a(x-d)^2) \tag{14}$$

$$y = \left(\frac{L}{1+\exp(-a_1(x-d_1))}\right) + \left(\frac{L}{1+\exp(a_2(x-d_2))}\right) - L \tag{15}$$

#### 3.6. Analysis 2 (dataset: COVID-19)[36]

**1) Dataset search and data review**

c. Dataset Search

To apply our idea to COVID-19 problems, one of the authors (KK) searched hyperbolic patterns using the search service Google (https://www.google.com/) provided by Google Inc., which allows displaying search results as "images." The search keyword was "COVID-19 AND Meta-analysis". In the case of displaying bubble charts instead of the error bars, the size of the bubble chart (inversely proportional to the length of the error bars) was converted in mind. Consequently, we selected a study reported by Matsushita et al (Matsushita, K. et al., *Glob Heart.* 2020;15(1):64.)[36] that included eight research papers in Figure 5[52-58,68].

d. Data review

As in the case of Analysis 1, we reviewed to evaluate numerical accuracy and appropriateness from the viewpoints of biomedicine. Since Matsushita et al[36] originally made web Figure 5 and excluded 17 studies [37,69-84] from avoiding duplication of studies in Wuhan city in the making of Figure 5, we inspected both of studies in Figure 5 (8 studies) and only in web Figure 5 (17 studies).

Based on the results shown below, considering the issue of comparability, we excluded the data reported by Yuan et al[68] and Wang L. et al[56]. Also, we re-calculated the age difference using data reported by Guan et al[54] **(see Fig. 2).**

As a side note, the numbers assigned to each point in figure 3 were the same numbers described in the original figure by Matsushita et al[36], and the correspondence relationship is the following **(Fig. S12)**: **No.2**: Cao et al (Cao, J. et al., *Intensive Care Med.* 2020;46(5):851-853.)[52], **No.7**: Deng et al (Deng, Y. et al., *Chin Med J (Engl).* 2020;133(11):1261-1267.)[53], **No.8**: Guan et al (Guan, W.J. et al., *N Engl J Med.* 2020;382(18):1708-1720.)[54], **No.19**: Wang D. et al (Wang, D. et al., *JAMA.* 2020;323(11):1061-1069.)[55], **No.20**: Wang L. et al (Wang, L. et al., *J Infect.* 2020;80(6):639-645.)[56], **No.21**: Wu et al (Wu, C. et al., *JAMA Intern Med.* 2020;180(7):934-943.)[57], **No.23**: Yuan et al

(Yuan, M. et al., *PLoS One.* 2020;15(3):e0230548)[68], **No.25**: Zhou et al (Zhou, F. et al., *Lancet.* 2020;395(10229):1054-1062.)[58].

(i) No.8 Guan et al (Guan, W.J. et al., *N Engl J Med.* 2020;382(18):1708-1720.)[54]

Matsushita et al[36] did not use the data divided into the severe and non-severe groups by Guan et al[54] but used the data divided into yes and no, using "Presence of Primary Composite End Point," which means entry to the intensive care unit (ICU), use of mechanical ventilation, or death.

Since Cao et al (Wuhan University Zhongnan Hospital in Wuhan; affiliation of Dr. Jianlei Cao: Department of Cardiology)[52] and Wang et al (Zhongnan Hospital of Wuhan University in Wuhan; affiliation of Dawei Wang, MD: Department of Critical Care Medicine)[55] also used ICU admission as a criterion for severe or non-severe, we examined the rate of severely ill patients and resulted in 21.4% (18/84) and 35.3% (36/102), respectively. However, in the case of using the "Presence of Primary Composite End Point," the percentage was only 6.5% (67/1032). Whereas, in the original categorization by Guan et al[54], the percentage was 18.7% (173/926). Therefore, we prioritized Guan et al's original classification of severe or non-severe[54].

(ii) No. 9 Guo et al (Guo et al. *JAMA Cardiol.* 2020;5(7):811- 818)[37] (only in eFigure5)

Three subgroup patterns appeared in a figure reported by Guo et al, although they did not mention it.

(iii) No. 20 Wang L. et al (Wang, D. et al., *JAMA.* 2020;323(11):1061-1069.)[56]

It was found that a significant matter of consideration on eligibility, the patient population reported by Wang L. et al was limited to over age 60[56]. The title was "Coronavirus disease 2019 in elderly patients: Characteristics and prognostic factors based on 4-week follow-up".

(iv) No.23 Yuan M. et al (Yuan M. et al., *PLoS One.* 2020;15(3):e0230548.)[68]

The zero cells appeared in the study reported by Yuan et al[68]. The difficulty of patient enrollment might cause a small sample size, which seemed to be a concern from the viewpoint of comparability (c.f., Guan W. et al, n=1099[54]; Zhou F. et al, n=191[58]; Wang D. et al, n=138[55]; Wu C. et al, n=201[57]; Cao J. et al, n=102[52]; Deng Y. et al, n=225[53]).

(v) The term "Cardiovascular disease (CVD)."

There was an inconsistency in the studies on "Cardiovascular disease (CVD)." For example, vascular diseases such as arrhythmia and arteriosclerosis are also classified as CVD, but in the studies reported by Guan et al[54] and Zhou et al[58], the term "Coronary heart disease" was used. Also, "Cardiac disease" was used by Yuan et al[68], "Heart disease" was used by Deng et al[53], and "Cardiovascular disease" was used by Wang L et al[56].

**2) Regression analysis**

Considering the problem of comparability, we re-calculated OR and visually grouped it into two hyperbolic patterns. In the case of S-shaped curve fitting, since there were many unknown coefficients for the number of data (3 unknown coefficients of $K_1$, $K_2$, and M), the regression analysis was performed after setting the zero point value.

In the fitting of upper and lower curves, since there were many unknown coefficients ($K_1$, $K_2$, M, a, b, c, see equation (7) & (8)), we first obtained the coefficient of M by the curve fitting of the S-shaped curve, and then performed curve fitting of parabolas. After substituting M for the x value of apex in equation for standard form of quadratic function (see equation (16) & (17)), regression analysis was performed on the data in the middle row of the figure **(Fig. S11, d-f)**. Finally, the S-shaped curve and the parabola were merged **(Fig. S11, g-i)**.

$$ax^2 + bx + c = a\left(x + \frac{b}{2a}\right)^2 - \frac{b^2-4ac}{4a} \quad (16)$$

$$-\frac{b}{2a} = M \quad (17)$$

**3) Calculation of weighted average**

In the earlier days group, the median age and the number of cases are tabulated by severe and non-severe cases as follows. Guan W. et al (severe n=173 [age: 52] vs. non-severe n=926 [age: 45])[54], Zhou F. et al (non-survival n=54 [age: 69] vs. survival n=137 [age: 52])[58], Wang D. et al. (ICU n=36 [age: 66] vs. non-ICU n=102 [age: 51])[55], Wu C. et al (ARDS n=84 [age: 58.5] vs. non-ARDS n=117 [age: 48])[57], and the whole of earlier days group (severe n = 347 vs. non-severe n = 1282).

The weighted average of severe and non-severe in the earlier days group was calculated from these values by the following formulas. In the earlier days group, the weighted average of severe and non-severe were 57.7 and 46.5, respectively.

$$Number\ of\ patients\ in\ earlier\ days\ group\ (severe) = 173 + 54 + 36 + 84 = \mathbf{347}$$

$$Weighted\ mean\ of\ earlier\ days\ group\ (severe)$$

$$= \frac{173}{\mathbf{347}} \times 52 + \frac{54}{\mathbf{347}} \times 69 + \frac{36}{\mathbf{347}} \times 66 + \frac{84}{\mathbf{347}} \times 58.5 \cong 57.7$$

$$Number\ of\ patients\ in\ earlier\ days\ group(non-severe) = 926 + 137 + 102 + 117$$

$$= \mathbf{1282}$$

$$Weighted\ mean\ of\ earlier\ days\ group\ (non-severe)$$

$$= \frac{926}{\mathbf{1282}} \times 45 + \frac{137}{\mathbf{1282}} \times 52 + \frac{102}{\mathbf{1282}} \times 51 + \frac{117}{\mathbf{1282}} \times 48 \cong 46.5$$

In the later days group, the median age and the number of cases are tabulated by severe and non-severe cases as follows. Cao J. et al (ICU n=18 [age: 66] vs. non-ICU n=84 [age: 31])[52], Deng Y. et al. (Death n=109 [age: 69] vs. survival n=116 [age: 48])[53], and the whole of later days group (severe n = 127 vs. non-severe n = 200).

The weighted average of severe and non-severe in the late-date group was calculated using the following formulas. In the late date group, the weighted average of severe and non-severe were 68.6 years and 40.9, respectively.

$$Number\ of\ patients\ in\ late\ date\ group\ (severe) = 18 + 109 = \mathbf{127}$$

$$Weighted\ mean\ of\ late\ date\ group\ (severe) = \frac{18}{\mathbf{127}} \times 66 + \frac{109}{\mathbf{127}} \times 69 \cong 68.6$$

$$Number\ of\ patients\ in\ late\ date\ group\ (non - severe) = 84 + 116 = \mathbf{200}$$

$$Weighted\ mean\ of\ late\ date\ group\ (non - severe) = \frac{84}{\mathbf{200}} \times 31 + \frac{116}{\mathbf{200}} \times 48 \cong 40.9$$

**4) Additional analysis: Regression analysis on the parabolic cylinder**

One of the authors (KK) found a way to fit an appropriate curve to the data reported by Guo et al, performing trial and error with his mathematical intuition **(Fig. 6)**. Firstly, he calculated the center of gravity of the data by each subgroup cluster (center of gravity: the average of the values on the horizontal axis x and the average of the values on the vertical axis y). Second, he obtained equations of three straight lines passing through the origin and centers of gravity. Thirdly, he obtained the equation of a straight line passing through each center of gravity and intersecting the straight lines obtained above. Fourthly, he re-set new origin as each center of gravity and regarded the above two crossed lines as a small cartesian coordinate system. Finally, he applied parabola fitting with Excel®. In this curve fitting, he used the data of distance between each data point and the straight line obtained secondary, and the data of distance between each data point and the straight line obtained firstly (see "distance from a point to a line" in a high school textbook).

Supplementary, our result suggested that the treatment of implicit function, which is unfamiliar in the medical and biological fields, is required. In this research, we used our method to fit the curves. However, a Fortran program created by Dr. Timmes[85], a researcher in astronomy, may also be helpful for future research.

**5) Making example data**

To explain the misuse of linear regression analysis in Guo et al[37], we made the following data to show the example. It allows being used in R by copying and pasting the following.

```
Value_X<-
c(0.04 ,0.08 ,0.12 ,0.16 ,0.2 ,0.24 ,0.28 ,0.32 ,0.36 ,0.4 ,0.44 ,0.48 ,0.52 ,0.56 ,0.6 ,0.64 ,0.68 ,0.72 ,0.7
6 ,0.8 ,0.84 ,0.88 ,0.92 ,0.96 ,1 ,1.04 ,1.08 ,1.12 ,1.16 ,1.2 ,1.24 ,1.28 ,1.32 ,1.36 ,1.4 ,1.44 ,1.48 ,1.52 ,
1.56 ,1.6 ,1.64 ,1.68 ,1.72 ,1.76 ,1.8 ,1.84 ,1.88 ,1.92 ,1.96 ,2 ,2.04 ,2.08 ,2.12 ,2.16 ,2.2 ,2.24 ,2.28 ,2.
32 ,2.36 ,2.4 ,2.44 ,2.48 ,2.52 ,2.56 ,2.6 ,2.64 ,2.68 ,2.72 ,2.76 ,2.8 ,2.84 ,2.88 ,2.92 ,2.96 ,3 ,3.04 ,3.08
,3.12 ,3.16 ,3.2 ,3.24 ,3.28 ,3.32 ,3.36 ,3.4 ,3.44 ,3.48 ,3.52 ,3.56 ,3.6 ,3.64 ,3.68 ,3.72 ,3.76 ,3.8 ,3.84 ,
3.88 ,3.92 ,3.96 ,4)

Value_Y<-
c(2.01742 ,2.02749 ,2.04454 ,2.02009 ,2.03445 ,2.04749 ,2.03641 ,1.99047 ,1.98671 ,2.06711 ,2.0977
2 ,2.00539 ,1.86985 ,2.01679 ,2.12183 ,1.94453 ,1.86497 ,1.87444 ,2.09483 ,1.91073 ,1.69244 ,1.709
68 ,1.81376 ,2.04219 ,1.69059 ,1.75939 ,1.95321 ,1.8417 ,1.58169 ,1.75585 ,1.73497 ,1.47847 ,1.911
5 ,1.44962 ,1.85063 ,1.32298 ,1.28437 ,1.74621 ,1.27232 ,1.32763 ,1.575 ,1.56609 ,1.5933 ,1.76707 ,
1.11264 ,1.06188 ,1.40139 ,0.94084 ,1.0756 ,1.38507 ,1.33408 ,1.54375 ,1.60257 ,1.02029 ,0.98612 ,
1.79094 ,0.97916 ,0.81212 ,1.1484 ,1.5168 ,1.70236 ,1.38945 ,1.69072 ,1.75042 ,1.67571 ,1.38623 ,1.
81503 ,1.80665 ,1.41073 ,2.3175 ,2.24852 ,1.7595 ,2.81818 ,1.93654 ,2.36998 ,2.0987 ,2.19539 ,2.44
747 ,2.57255 ,3.24637 ,2.78881 ,3.51638 ,2.68107 ,2.87259 ,4.3749 ,3.13393 ,3.92099 ,4.1223 ,3.785
84 ,4.9666 ,5.4888 ,5.70712 ,4.84752 ,5.1409 ,6.31637 ,5.53198 ,6.89257 ,7.86387 ,7.98237 ,8.64705
)
```

### 4. Supporting discussions

#### 4.1. Supporting discussion 1: history of traveler's thrombosis

In 2004, Adi et al performed a systematic review and meta-analysis on traveler's thrombosis. They reported that more than 8 hours of flight with other risk factors for thrombosis might be involved in the onset, but it was not conclusive evidence[2]. In 2007, Philbrick et al performed a systematic review study and reported that more than 6 hours of flight with other risk factors were involved in developing thrombosis[7]. In the same year, Spencer reviewed the article in the ACP Journal Club produced by the American College of Physicians and commented that more than 8 hours of flight would be a risk[8]. Also, Kuipers et al[6] performed a systematic review. They reported that flights of 4 hours or more were risk factors for symptomatic events, and flights of 12 hours or more were risk factors for pulmonary embolism (PE)[6].

However, in 2008, the discussion has returned to the beginning because Trujillo-Santos et al performed a systematic review & meta-analysis and reported that long-distance travel was a risk, but the relationship was weak[9].

In 2009, to find out the cause of the above, Chandra et al performed a meta-analysis (meta-regression analysis)[10] using a dataset derived from the data of 4 original studies[1,3,5,26], which were acquired by Martinelli et al, Cannegieter et al, Parkin et al, and Kuipers et al Chandra et al. They concluded that the cause of controversy was caused by “referred control,” which was a type of inappropriate selection of control[10].

Some researchers regarded that the study clarified a confusing body of evidence. Also, the study received positive feedback in the professional community[15,17], but some disagreement was also proposed. For example, in the editorial article, Vandenbroucke et al showed their argument that the discussion by Chandra et al was incomplete[11].

Moreover, in the subsequent articles, clear conclusions have not appeared[12-14,16]. Considering this situation, we thought that the possibility of some overlooked information should be considered.

### 4.2. Supporting discussion 2: research situations of COVID-19-related thrombosis

Historically, thrombosis research has a relatively long history in medicine. In the 19th century, Virchow's triad, named after Rudolf Virchow (1821-1902), had already been established. Virchow's triad contains three statements on the element of thrombus formation that is "Hypercoagulability," "Stasis," and "Endothelial injury"[18,86]. In 1948, when molecular biology was still in its early days (please imagine the DNA double helix model was proposed in 1953), blood coagulation factor 13 was already identified. So, the number of unsolved problems in coagulation physiology was relatively small than in immunology and neuroscience.

Nevertheless, it had already been known that forming microthrombus as a defense mechanism to trap pathogens such as viruses locally and disseminated intravascular coagulation (DIC) caused by confusion of the mechanism. However, in the above classical research field, some review articles informed that the problem of COVID-19 was not only a quantitatively challenging problem within the meaning that there were many infected patients, but also a problem that felt like a new experience to researchers as a reaction of the human body to infectious diseases. For example, Barrett et al mentioned that "Although DIC is a long-studied phenomenon, there has never been a disease like COVID-19 that so consistently causes thrombotic DIC in large numbers of patients"[19] as a comment on the interim guidance from the International Society for Thrombosis and Hemostasis[20].

Additionally, in articles related to COVID-19, Callaway et al mentioned in the title "Six months of coronavirus: the mysteries scientists are still racing to solve"[21]. Moreover, Tal et al mentioned in the title "Venous Thromboembolism Complicated With COVID-19: What Do We Know So Far?"[22], and Marietta et al mentioned in the title "COVID-19, coagulopathy and venous thromboembolism: more questions than answers "[23]. Thus, the words "mystery," "complexity," and "more questions than answers" appeared in the titles of these recent articles.

Indeed, the new coronavirus is a novel pathogen for humans, but there has been no change in the human body since the 19th century. Therefore, the above situation should also be suspected of some overlooked information, as in the case of the traveler's thrombosis.

This situation was similar to that of the traveler's thrombosis, and we started this analysis with the inspiration from the famous mathematician Polya who stated that solving a similar problem helped us solve a more complex problem[87].

### 4.3. Supporting discussion 3: two high-risk periods and two types of high-risk groups?

We obtained uniform results from two analyses in different datasets (Analysis 1A: 7.1 hours and 11.8 hours, Analysis 1B: 9.2 hours and 12.1 hours), gathered by Chandra et al[10] **(Fig. 3, b)** and Philbrick et al[7] **(Fig. S3, b)**. Also, there were two peaks at the time, around 7 hours and 12 hours, in the data reported by Clérel and Caillard **(Fig. S4)**. Therefore, we suspect two risk periods for the onset of thrombosis.

Also, we hypothesized two types of the group with a high risk of thromboses, such as high-risk group A and high-risk group B (risk of A > risk of B). Thus, high-risk group A develops PE in the first high-risk period. In contrast, high-risk group B passes the first risk time zone and develops DVT on a long flight (10 hours or more). Although this story did not explain why some high-risk group A present no DVT, considering a research report on the "factor V Leiden paradox," which reported the pathophysiology of PE and DVT was different [44], the pathophysiology of PE and DVT might be different in travel as well.

Additionally, in the result of the correlogram, the interval between the wave's peaks was about 18.35 hours, but the valley point was not the midpoint between the peaks. The length from the first peak (left peak) to the valley (minimum) and from the valley to the next peak (right peak) was 10.73 hours and 7.61 hours, respectively **(Fig. S4)**. This result may suggest complex periodic fluctuations. Those results matched the discussion on the above high-risk period, although this is a discussion about the same data because we use the same data for analysis by using a correlogram.

On the other hand, the time point divided equally between the two peaks (18.35 hours) was 9.18 hours, close to eight hours. This "eight hour" has been a component of the principle of "8 hours sleep, 8 hours work, 8 hours free time" since the labor movement in Chicago, USA, 1886, in human society. Also, the operation of the 8-hour principle is related to circadian rhythms (e.g., 8 hours of work during the day and 8 hours of sleep at night).

Also, the effects of circadian rhythms on the cardiovascular system are already known (e.g., morning surge). Therefore, it seems that the circadian rhythm underlies the periodic fluctuations observed on the correlogram, which has undergone some modification. Perhaps the circadian rhythm of the departure point and the circadian rhythm of the arrival point (Paris) are mixed, resulting in complex periodic fluctuations.

#### 4.4. Supporting discussion 4: for more discussion on Traveler's thrombosis in Women and Men

##### 1) Cyclic patterns newly founded in this research and OC

Despite the exposures in Cannegieter et al being more complex (air travel, train, bus, and car trip)[3] than Kelman et al (only air travel)[28], the wave in the data reported by Cannegieter et al. was clear, but the wave in the data reported by Kelman et al was not clear.

The data reported by Cannegieter et al contained many car trips (air travel, train, bus, and car trip). These cars might be honeymoon cars.

Kelman et al described that although the risk increased within 2 to 4 weeks of flight (hazard period), the risk was low after the period because travelers were healthier than the average of a group (healthy traveler effect)[28]. Also, Rothman mentioned the "persistence of travel-related risk" and "spline regression"[4]. They might realize the position around 3-4 weeks as a valley between two distributions (honestly, we thought the same as them before we learned the cyclic patterns by spending long-time severe observations and non-linear regression analysis).

##### 2) The difference in the cyclic patterns and HRT

We found a slowdown of the increase of thrombosis onset at Paris airports in 1998, the year of HERS result was issued **(Fig. S5)**. Also, our trend analysis showed that the number of literature on thrombosis decreased since around 2000 **(Fig. S6).** Moreover, our merged data from four studies[26,27,33,34] on PE from the 1970s to the 2000s showed that the rate of women was most pronounced during the late 1990s **(Table S3)**.

Although the existence of the women patients not "menopausal" stage women (40s to 50s) but among around 70-75 years of age seems to disagree with HRT use **(Fig. S4, d, & Fig. S8, d)**, the subjects of the WHI study were "postmenopausal" women (age 50-79) for prophylactic use for cardiovascular problems.

**3) Men's life stage and travel related phycological stress**

Every four studies[26,27,33,34] showed inconsistent results (correlated or inversely correlated). This situation seems to be the same as the parable of "blind men and an elephant." Considering the correlation between age and the risk of thrombosis, the above phenomena may be one of the causes of controversial discussions. Also, It seems worth noting that Grant commented on specific age group (50-59) in a rapid response to Kelman et al's work (electronic comment on BMJ website)[88], although the word was related to the context of OC and HRT.

**4) Changing the framework of the disease concept of "travel-related thrombosis."**

Although the results of a meta-regression analysis (Chandra et al's result[10] and our results of S-curve fittings) showing that the risk of thrombosis increases with travel duration have seemed to support the in-flight environment theory, the possibility may exist that what essentially correlates with the risk of thrombosis is "increased health risks with aging" or "psychological stress associated with rising job titles," since men's data shows an increase in travel time with age. Nemeth et al experimentally showed that psychological stress (fear caused by a horror movie) was associated with a rise in blood coagulant factor VIII[89].

**5) Controversial discussions and overlooked patterns**

Recently, we have thought that the concept of "medical research test" or "inspection of medical data analysis" can be developed analogically as an extension of the medical inspection, including laboratory tests for each patient for each patient. Like medical inspection using equipment, detecting overlayed hyperbolic shape patterns may be a tool or marker of some overlooked information **(Fig. S1, e)**.

### 4.5. Supporting discussion 5: linear regression analysis

Drawing regression line to the scatter plot seems inappropriate because it is not acceptable to suggest the validity of applying a linear regression analysis only by the significant result of a test for regression coefficient. Also, a statistical test result generally becomes significant more easily as the sample size increases.

Additionally, we found unrecognized patterns during the data review process in a figure reported by Guo et al[37], which showed a relationship between N-terminal pro-brain sodium diuretic peptide (NT-proBNP) and Troponin T (TnT). There were three clusters of subgroup patterns, and the third subgroup pattern was similar to the pattern of ST-elevating myocardial infarction in the figure report by Budnik et al (*Int J Cardiol.* 2016;15(219), Fig. 1) [38]. Also, this tilted parabola pattern has already appeared in Sugawa et al (*Sci Rep.* 2018;8(1), Figure 1) [41] and Satyan et al (*Am J Kidney Dis.* 2007;50(6), Figure 1)[40]. These indicated that this pattern was reproducible.

However, those patterns were not recognized by each author. In another paper, Sugawa divided the 2-dimensional scatters plot into four areas by crossed axis and compared each area (*J Med Diagn Meth.* 2017;6(2), Figure 1)[90]. Therefore, Sugawa may have noticed the existence of a parabola, although he did not entirely recognize it.

As a cause of the above-confused situation, it is conceived that many researchers applied linear regression analysis inappropriately by focusing only on statistical significance, such as "$p < 0.001$".

To explain the misuse of linear regression analysis, we made virtual data using random number generation in spreadsheet software **(see Supporting information: Method details)**. The following are estimated equations by the regression analysis and the statistical test result performed using a program on the statistical analysis software R platform **(Fig. S13, f)**.

In panel f of **Fig. S13**, the formula for the regression line is shown below. Also, the estimated equation is expressed in the following line. The test result is significant due to a large sample (n = 100).

$$y = \beta_0 + \beta_1 x$$

$$y = 0.7280^{**} + 0.8313^{***}x$$

significance codes: '***' 0.001 '**' 0.01 '*' 0.05 '.' 0.1 ' ' 1

In this panel, the formula for the regression curve is shown below. Also, the estimated equation is expressed in the next line.

$$y = \beta_0 + \beta_1 x + \beta_2 x^2 + \beta_3 x^3 + \beta_4 x^4$$

$$y = 2.14594^{***} - 0.54052\, x + 0.49655\, x^2 - 0.42523^{\cdot}\ x^3 + 0.10680^{***} x^4$$

significance codes: '***' 0.001 '**' 0.01 '*' 0.05 '.' 0.1 ' ' 1

A statistical test on a coefficient of line or curve is helpful for variable selection if there are many variables and visual confirmation is impossible (e.g., multivariate regression analysis). However, significance does not indicate judgment to remove coefficients, but the p-value relatively shows the importance of the coefficients. In this equation, although the p-value indicates non-significant, excluding coefficients ("-0.54052", "0.49655", and "-0.42523") results in a poor curve fitting the data **(dashed line in Fig. S13, f)**.
